# Efficacy of antioxidant therapy in mild to moderate SARS-CoV-2 infection: A pilot experimental arm of the CanTreatCOVID adaptive platform trial

**DOI:** 10.64898/2026.08.21.26360704

**Authors:** Banafshe Hosseini, David Jenkins, Peter Daley, Kerry McBrien, Srinivas Murthy, Amanda Condon, Bruno R. da Costa, Michelle Greiver, Peter Jüni, Peter Selby, Norman Umali, Meichen Liu, Haolun Shi, Kawsika Sivayoganathan, Darshna Patel, Melanie Paquette, Helen Hue My Nguyen, Marc Malty, Aleksandra Nedeljkovic, Nerissa Situ, Gabriel So, Ethan Belo, Yueyang Han, Andrew D. Pinto

**Author notes:** Address correspondence to: Banafshe (Benita) Hosseini, Upstream Lab, MAP Centre for Urban Health Solutions, Li Ka Shing Knowledge Institute, Unity Health Toronto 30 Bond Street, Toronto, ON, M5B 1W8.

## Abstract

**Background:** Although the acute phase of the COVID-19 pandemic has passed, SARS-CoV-2 continues to cause outpatient morbidity. Antioxidant micronutrients support immune regulation and may offer a low-cost, scalable adjunctive treatment in early infection.

**Objective:** To evaluate a pilot combination antioxidant therapy within CanTreatCOVID.

**Methods:** This pilot sub-protocol enrolled non-hospitalized adults across five Canadian provinces (September 5^th^, 2024-March 31^st^, 2025) with mild-to-moderate SARS-CoV-2 infection within five days of symptom onset. Participants were randomized to usual care plus a 10-day antioxidant regimen (selenium 300 µg, zinc 40 mg, lycopene 45 mg, vitamin C 1.5 g) or usual care alone. Pilot objectives assessed feasibility, retention, adherence, and safety. The primary outcome was hospitalization or death within 28 days; exploratory outcomes included recovery and symptom measures by day 14.

**Results:** Eighty-one participants were randomized (41 antioxidant; 40 usual care). Retention was high 85.4% antioxidant; 82.5% usual care), and 90.2% of antioxidant participants completed the intervention course. Adverse events were infrequent (9.8% vs 2.5%), with no serious adverse events reported. No deaths occurred in either group; no hospitalizations occurred in the antioxidant arm versus 2/40 (5%) in usual care. By day 14, recovery was reported in 32/40 (80.0%) participants receiving antioxidants versus 23/36 (63.9%) in usual care (OR 2.128; 95% CI 0.747–4.871). Sustained alleviation of all symptoms occurred in 38/40 (95.0%) versus 29/36 (80.6%), respectively (OR 3.498; 95% CI 0.872–10.017). Return to usual activity by day 14 occurred in 38/40 (95.0%) versus 30/36 (83.3%) (OR 3.113; 95% CI 0.762–9.022). Adjusted between-group differences in dietary intake were not statistically significant.

**Conclusions:** Combination antioxidant therapy was feasible to deliver in a decentralized outpatient setting, with high adherence and tolerability. While the trial was not powered for definitive efficacy conclusions, consistent directional improvements across symptom outcomes support evaluation of this host-directed antioxidant strategy in larger trials.

**Trial registration number:** https://clinicaltrials.gov/study/NCT05614349

## Introduction

Although the acute phase of the COVID-19 pandemic has passed, SARS-CoV-2 continues to circulate globally and remains a cause of outpatient morbidity, particularly among older adults and individuals with comorbidities.^1,2^ As clinical care has shifted from crisis response to sustainable long-term management, there is ongoing interest in identifying low-cost, scalable therapies that can be deployed early in the course of infection. While direct-acting antivirals have demonstrated benefit in selected high-risk populations^3^ complementary strategies that target host responses may still have value, especially in community settings.

Antioxidants are compounds that neutralize reactive oxygen species and support the body’s endogenous defense systems^4^. In the context of viral respiratory infections, they have been proposed as adjunctive therapies to mitigate oxidative stress and downstream inflammatory injury.^5^ Viral infections induce oxidative stress by producing reactive oxygen species in host cells, which, if not counterbalanced by antioxidant defense mechanisms, can lead to oxidative stress and the emergence of more virulent strains.^6,7^ The depletion of antiviral defenses and elevated production of inflammatory cytokines have been identified as key factors in SARS-CoV-2 infection.^5,8^ Modulating redox balance therefore represents a biologically plausible adjunctive strategy^5^, particularly in mild to moderate cases where preventing escalation is the primary goal.

Several systematic reviews have suggested that antioxidants may have potential benefits for COVID-related outcomes, including preventing disease progression in patients with higher levels of antioxidants.^9,10^ Our team recently conducted a systematic review and meta-analysis of randomized controlled trials on antioxidant therapies in SARS-CoV-2 infection synthesized evidence across interventions including zinc, vitamin C, vitamin A, and combination regimens.^11^ While results were mixed and definitive conclusions about clinical efficacy were limited by heterogeneity and risk of bias, our analysis identified signals of potential benefit in symptom resolution, inflammatory modulation, and hospitalization outcomes that warrant further investigation.^11^

Rather than focusing on a single micronutrient, we hypothesized that a combination strategy targeting complementary aspects of the antioxidant defense system might offer greater biological impact. The endogenous antioxidant system depends on adequate micronutrient availability to support enzymatic pathways such as superoxide dismutase and glutathione peroxidase, while exogenous antioxidants can directly neutralize reactive oxygen species^12^. A combined approach may therefore strengthen intrinsic defenses while providing additional redox buffering during acute infection. To explore this hypothesis, we introduced a pilot experimental sub-protocol within the CanTreatCOVID adaptive platform trial^13^. This arm evaluated a 10-day course of combination antioxidant therapy, comprising selenium (300 µg), zinc (40 mg), lycopene (45 mg), and vitamin C (1.5 g), compared with usual care among non-hospitalized patients with mild to moderate SARS-CoV-2 infection. The aim of this paper is to report the 28- day clinical outcomes from this pilot antioxidant arm, specifically examining symptom duration, symptom severity, and health service use in the first 28 days following randomization.

## Methods

### Study Design

This study was conducted as a pilot experimental sub-protocol within the CanTreatCOVID adaptive platform trial, a multi-provincial, individually randomized outpatient trial. The CanTreatCOVID trial is registered at ClinicalTrials.gov (NCT05614349), and the full platform protocol has been published previously^13^. Several deviations from the trial protocol occurred in this pilot sub-study. These included informing participants of early study closure, delivery of study medication outside the protocol-specified time window, and completion of follow-up assessments outside the planned follow-up window. In addition, some data were incomplete or missing, and some follow-up assessments were only partially completed due to participants being unreachable despite repeated contact attempts.

The trial was approved by Research Ethics Boards in five Canadian provinces: Unity Health Toronto Research Ethics Board – St. Michael’s Hospital (Ontario), the Conjoint Health Research Ethics Board, University of Calgary (Alberta), the University of Manitoba Biomedical Research Board - Research Ethics Bannatyne (Manitoba), the University British Columbia Children’s & Women’s Research Ethics Board (British Columbia), and the Newfoundland and Labrador Health Research Ethics Board (Newfoundland and Labrador).

CanTreatCOVID was designed to evaluate therapeutic strategies for non-hospitalized adults with mild to moderate SARS-CoV-2 infection. At the time of this sub-protocol, active arms included usual care alone and usual care plus the antioxidant therapy. The adaptive structure allowed new arms to be added or discontinued based on interim review by an independent Data and Safety Monitoring Committee (DSMC) and recommendations from the Canadian COVID-19 Out-Patient Therapeutics Committee. The antioxidant arm was introduced as a pilot arm within this platform.

### Participants

Participants were eligible for this sub-protocol if they met all platform-level inclusion criteria and none of the platform-level exclusion criteria. Platform inclusion criteria included age≥ 50 years, or age 18-49 years with at least one high-risk chronic medication condition or immunosuppression, and symptom onset within 5 days prior to enrollment. Platform exclusion criteria included current hospitalization or emergency department stay >24 hours, prior randomization in the platform, enrollment in incompatible therapeutic trials, contraindications to study treatments, or inability to provide informed consent.

Additional exclusions specific to the antioxidant arm included age <19 years, known or suspected pregnancy or breastfeeding, allergy or intolerance to study supplements, advanced chronic kidney disease (CKD stage 3: eGFR ≥30 to <60 mL/min, and severe renal impairment (eGFR <30 ml/min, CKD stage 4-5), liver disease awaiting transplantation, history of calcium oxalate kidney stones, recent head and neck cancer (within 5 years), warfarin use, baseline use of selenium (≥300 µg/day), zinc (≥40 mg/day), lycopene (≥45 mg/day), or vitamin C (≥1500 mg/day), and consumption of omega-3 fatty acid supplements at baseline with unwillingness to discontinue during the intervention period. Consent was obtained through e-consent.

### Intervention

Participants randomized to the antioxidant arm received usual care according to provincial guidance plus a 10-day course of combination antioxidant therapy consisting of selenium (300 µg), zinc (40 mg), lycopene (45 mg), and vitamin C (1.5 g). Supplements were administered as three tablets once daily and initiated within 5 days of symptom onset. Supplements were couriered directly to participants’ homes through provincial research hubs. Participants were instructed to complete the full 10-day course. If hospitalization occurred after initiation, continuation was at the discretion of the treating clinician. Compliance was assessed through self-reported daily diaries for 14 days following randomization.

Supplements were manufactured in Canada by Advanced Orthomolecular Research (AOR, Calgary, Canada).

### Randomization

Randomization occurred at enrollment through a centralized web-based system maintained by the Applied Health Research Centre (Unity Health Toronto). Participants were randomized in equal allocation ratios across all eligible arms at the time of enrollment. Randomization was stratified by age (<65 years vs ≥65 years).

### Blinding and Bias Mitigation

The trial was open label; participants and recruiting clinicians were aware of treatment allocation. However, outcome analysis was conducted by investigators blinded to allocation. Furthermore, steering Committee members did not have access to unblinded comparative data. To minimize attrition bias, participants were compensated for completing study encounters regardless of adherence. Data on co-interventions and contamination (receipt of study therapeutics outside the trial) were collected.

### Recruitment

Participants were recruited through primary care practices, emergency departments, outpatient clinics, community outreach, and public-facing materials, with the option to enroll by contacting the study’s toll-free hotline directly. The trial was fully decentralized, with no in-person study visits required; participants could be recruited from anywhere within the five participating provinces. Screening was conducted in three sequential steps: an initial eligibility assessment by a research assistant, followed by a medication and contraindication review by a study pharmacist, and final confirmation of eligibility by the site investigator prior to randomization. Recruitment started on different days across the five participating provinces: September 5, 2024 (Newfoundland and Labrador), September 11, 2024 (Alberta), September 26, 2024 (Manitoba), October 22, 2024 (British Columbia), November 20, 2024 (Ontario). Recruitment ended on March 31^st^, 2025, across all sites.

### Outcomes

The platform-level primary outcome for CanTreatCOVID was all-cause hospitalization or death within 28 days of randomization. Given the small sample size and low expected event rate in this outpatient pilot cohort, this sub-protocol was not powered to detect differences in severe outcomes. Accordingly, the primary objectives of this pilot were to assess feasibility, including recruitment, retention, adherence to the intervention, and safety.

Exploratory clinical outcomes focused on participant-reported recovery and symptom trajectories over the first 28 days following randomization. Recovery and symptom outcomes were collected using daily electronic diaries from day 1 to day 14, with additional follow-up assessments at days 21 and 28. Recovery was assessed using the question: “Do you feel recovered today? i.e., symptoms associated with illness are no longer a problem)”, consistent with measures used in the PRINCIPLE^14^ and PANORAMIC^3^ trials. Symptom severity was assessed daily using the questions: “How well are you feeling today? Please rate how you are feeling now using a global rating scale (1 = no symptoms to 4 = very severe symptoms), alongside symptom-specific ratings categorized as no problem, mild, moderate, or major.

Prespecified exploratory outcomes included recovery by day 14; complete alleviation of all symptoms by day 14; sustained alleviation of symptoms, defined as the first of three consecutive days of symptom resolution or only mild symptoms without subsequent relapse by day 28; return to usual health; and return to usual activity. Return to usual health and activity were assessed using items derived from the FluPro Plus questionnaire^15^;

### Data collection

Data were collected using REDCap® hosted on secure servers at Unity Health Toronto. At baseline, participants provided sociodemographic data (age, sex assigned at birth, gender identity, education, income, ethnicity, rurality), medical history, vaccination status, and concomitant medications. Participants completed daily electronic diaries for 14 days, followed by online surveys at days 21 and 28. Participants without internet access were contacted by telephone to complete study assessments. Study staff attempted follow-up after two consecutive missed diary entries prior to day 7 or day 14. Participants received $30 CAD for each completed study encounter.

### Dietary assessment

To characterize baseline dietary antioxidant intake and assess potential changes during the intervention period, we obtained two 24-hour dietary recalls (24-HDRs) from participants randomized to the antioxidant therapy arm and to usual care. One recall was conducted at baseline and a second at the end of the 10-day intervention period. The 24-HDRs were conducted by telephone by trained interviewers. During each recall, participants were asked to describe in detail all foods and beverages consumed in the preceding 24 hours. Recalls were scheduled to capture both a weekday and a weekend day (Saturday or Sunday), given known differences in dietary patterns and alcohol consumption between weekdays and weekends. The recall period was defined as the 24 consecutive hours between midnight on day one and midnight on the following day. To improve portion size estimation, participants were encouraged to use measuring cups and measuring spoons during the telephone interview. Across the study, each participant completed a total of four 24-HDRs (two at baseline and two at day 10).

### Statistical analysis

Analyses for efficacy parameters were conducted using an intention-to-treat (ITT) population, defined as all randomized patients. Dietary intake analyses were conducted among participants with complete 24-hour dietary recall data at baseline and day 10. For each nutrient, within-group changes from baseline to day 10 were assessed using paired t-tests. Mean change values with 95% confidence intervals (CI) were calculated as day 10 minus baseline intake. Between-group differences in change were estimated using linear regression models with change in nutrient intake as the dependent variable and treatment group as the independent variable, adjusting for baseline nutrient intake. Regression coefficients (β) with 95% CIs were reported. A total of 61 nutrients were examined.

Clinical outcomes were analyzed using a Bayesian framework consistent with the CanTreatCOVID platform methodology^13^. For binary outcomes, including hospitalization or death by day 28, recovery by day 14, complete alleviation of symptoms, sustained symptom alleviation, return to usual health, and return to usual activity, Bayesian logistic regression models were used to estimate odds ratios with 95% credible intervals and the posterior probability of superiority of antioxidant therapy compared with usual care. Covariates included age, vaccination status, comorbidity, and receipt of nirmatrelvir/ritonavir (Paxlovid) during the intervention, as this antiviral was part of usual care in Canada for higher-risk patients (e.g., ≥65 years) during the study period.

Safety outcomes were summarized descriptively as the number of adverse events and the number of participants experiencing at least one event in each treatment group. All statistical analyses were conducted using Python 3.12.11. Statistical significance for exploratory analyses was defined as a two-sided p value <0.05.

## Results

### Participant flow

**Figure 1** presents the CONSORT flow diagram. A total of 165 individuals were screened for eligibility. Of these, 84 were deemed ineligible. The most common reasons for ineligibility included symptom onset more than five days prior to screening or absence of a confirmed SARS-CoV-2 test (n = 21), medical contraindications (n = 18), current use of antioxidant supplements above protocol-specified doses (n = 11), age 18–49 years without a qualifying chronic condition (n = 6), and inability to provide informed consent (n = 28).

**Figure 1.**
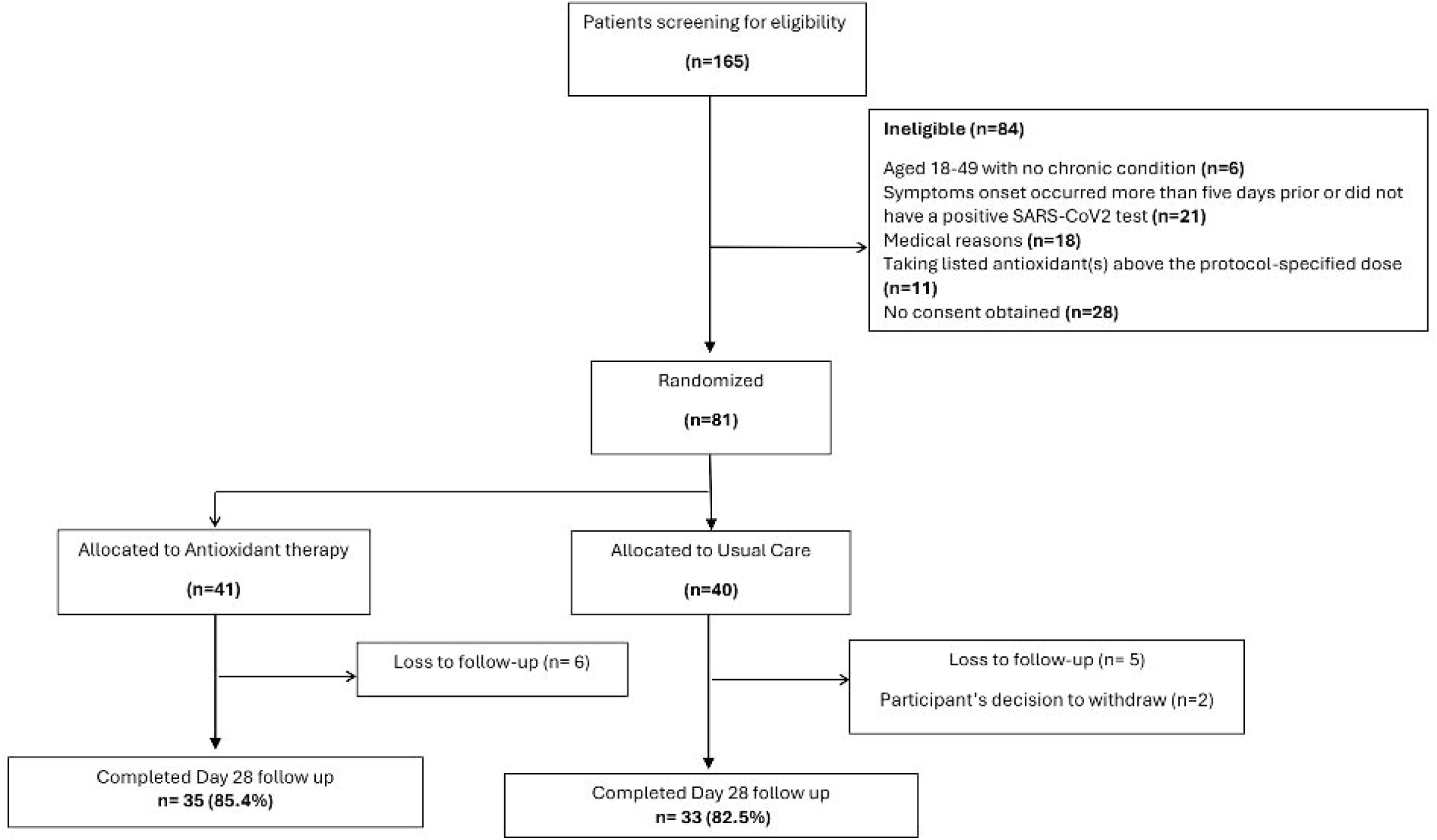
CONSORT flow diagram of participant screening, randomization, and follow-up in the antioxidant therapy pilot sub-protocol of the CanTreatCOVID adaptive platform trial.

### Baseline characteristics

Baseline characteristics are presented in Table 1 and were comparable between groups. The mean age was 57.4 years (SD 12.7) in the antioxidant arm and 57.6 years (SD 13.4) in usual care. In the antioxidant arm, 13/41 (31.7%) were male compared with 5/40 (12.5%) in usual care. Most participants self-identified as White (30/41, 73.2% vs 24/35, 68.6%) in the antioxidant and usual care arms, respectively.

**Table 1:** Baseline characteristics of study participants.

|  |  | **Antioxidant therapy (N=41)** | **Control (N=40)** |
| --- | --- | --- | --- |
| **Age, mean(SD) [min,max]** |  | 57.4 (12.7)<br>[22,77] | 57.6 (13.4)<br>[25,82] |
| **Sex, n(%)** |  |  |  |
|  | *Male* | 13 (31.7%) | 5 (12.5%) |
|  | *Female* | 28 (68.3%) | 30 (75.0%) |
|  | *Intersex* | 0 (0.0%) | 0 (0.0%) |
|  | *Missing* | 0 (0.0%) | 5 (12.5%) |
| **Ethnicity category, n(%)** |  |  |  |
|  | *White* | 30 (73.2%) | 24 (68.6%) |
|  | *Asian* | 6 (14.6%) | 9 (25.7%) |
|  | *Black* | 0 (0.0%) | 0 (0.0%) |
|  | *Indigenous* | 1 (2.4%) | 1 (2.9%) |
|  | *Mixed Race* | 4 (9.8%) | 1 (2.9%) |
|  | *Other* | 0 (0.0%) | 0 (0.0%) |
|  | *Missing* | 0 (0.0%) | 5 (12.5%) |
| **Duration symptoms at baseline in days, mean(SD)** |  | 2.7 (1.1) | 2.7 (1.3) |
| **Duration symptoms at baseline in days, median(IQR)** |  | 3.0 (2.0,3.0) | 3.0 (2.0,4.0) |
| **Number of vaccine doses, n(%)** |  |  |  |
|  | *None* | 0 (0.0%) | 1 (2.6%) |
|  | *Less than 2* | 0 (0.0%) | 0 (0.0%) |
|  | *2 or more* | 41 (100.0%) | 37 (97.4%) |
|  | *Missing, n(%)* | 0 (0.0%) | 2 (5.0%) |
| **Body Mass Index, median(IQR)** |  | 27.6 (23.8,32.0) | 24.8 (23.0,30.2) |
| **Baseline symptoms** |  |  |  |
| **Fever, n(%)** |  |  |  |
|  | *No problem* | 21 (51.2%) | 16 (45.7%) |
|  | *Mild problem* | 11 (26.8%) | 16 (45.7%) |
|  | *Moderate problem* | 8 (19.5%) | 2 (5.7%) |
|  | *Major problem* | 1 (2.4%) | 1 (2.9%) |
|  | *Missing, n(%)* | 0 (0.0%) | 5 (12.5%) |
| **Cough, n(%)** |  |  |  |
|  | *No problem* | 0 (0.0%) | 5 (14.3%) |
|  | *Mild problem* | 22 (62.9%) | 13 (37.1%) |
|  | *Moderate problem* | 9 (25.7%) | 9 (25.7%) |
|  | *Major problem* | 4 (11.4%) | 8 (22.9%) |
|  | *Missing, n(%)* | 0 (0.0%) | 5 (12.5%) |
| **Shortness of breath, n(%)** |  |  |  |
|  | *No problem* | 29 (70.7%) | 19 (54.3%) |
|  | *Mild problem* | 8 (19.5%) | 11 (31.4%) |
|  | *Moderate problem* | 4 (9.8%) | 3 (8.6%) |
|  | *Major problem* | 0 (0.0%) | 2 (5.7%) |
|  | *Missing, n(%)* | 0 (0.0%) | 5 (12.5%) |
| **Loss of smell or taste, n(%)** |  |  |  |
|  | *No problem* | 25 (61.0%) | 18 (51.4%) |
|  | *Mild problem* | 15 (36.6%) | 12 (34.3%) |
|  | *Moderate problem* | 0 (0.0%) | 5 (14.3%) |
|  | *Major problem* | 1 (2.4%) | 0 (0.0%) |
|  | *Missing, n(%)* | 0 (0.0%) | 5 (12.5%) |
| **Muscle ache, n(%)** |  |  |  |
|  | *No problem* | 14 (34.1%) | 10 (28.6%) |
|  | *Mild problem* | 15 (36.6%) | 10 (28.6%) |
|  | *Moderate problem* | 11 (26.8%) | 10 (28.6%) |
|  | *Major problem* | 1 (2.4%) | 5 (14.3%) |
|  | *Missing, n(%)* | 0 (0.0%) | 5 (12.5%) |
| **Nausea/ Vomiting, n(%)** |  |  |  |
|  | *No problem* | 35 (85.4%) | 28 (80.0%) |
|  | *Mild problem* | 4 (9.8%) | 6 (17.1%) |
|  | *Moderate problem* | 2 (4.9%) | 1 (2.9%) |
|  | *Major problem* | 0 (0.0%) | 0 (0.0%) |
|  | *Missing, n(%)* | 0 (0.0%) | 5 (12.5%) |
| **Fatigue, n(%)** | *No problem* | 2 (4.9%) | 5 (14.3%) |
|  | *Mild problem* | 15 (36.6%) | 7 (20.0%) |
|  | *Moderate problem* | 15 (36.6%) | 12 (34.3%) |
|  | *Major problem* | 9 (22.0%) | 11 (31.4%) |
|  | *Missing, n(%)* | 0 (0.0%) | 5 (12.5%) |
| **Difficult concentrating, n(%)** | *No problem* | 12 (29.3%) | 11 (31.4%) |
|  | *Mild problem* | 18 (43.9%) | 14 (40.0%) |
|  | *Moderate problem* | 9 (22.0%) | 6 (17.1%) |
|  | *Major problem* | 2 (4.9%) | 4 (11.4%) |
|  | *Missing, n(%)* | 0 (0.0%) | 5 (12.5%) |
| **Anxious mood, n(%)** | *No problem* | 28 (68.3%) | 20 (57.1%) |
|  | *Mild problem* | 9 (22.0%) | 8 (22.9%) |
|  | *Moderate problem* | 3 (7.3%) | 6 (17.1%) |
|  | *Major problem* | 1 (2.4%) | 1 (2.9%) |
|  | *Missing, n(%)* | 0 (0.0%) | 5 (12.5%) |
| **Any symptom rated moderate or major, n(%)** |  | 32 (78.0%) | 28 (80.0%) |
|  | *Missing, n(%)* | 0 (0.0%) | 5 (12.5%) |
| **Comorbidities** |  |  |  |
| **Lung disease, n(%)** |  | 6 (14.6%) | 10 (25.0%) |
|  | *Missing, n(%)* | 0 (0.0%) | 0 (0.0%) |
| **Heart disease, n(%)** |  | 8 (19.5%) | 9 (22.5%) |
|  | *Missing, n(%)* | 0 (0.0%) | 0 (0.0%) |
| **Kidney disease, n(%)** |  | 0 (0.0%) | 0 (0.0%) |
|  | *Missing, n(%)* | 0 (0.0%) | 2 (5.0%) |
| **Liver disease, n(%)** |  | 1 (2.4%) | 0 (0.0%) |
|  | *Missing, n(%)* | 0 (0.0%) | 0 (0.0%) |
| **Neurological disease, n(%)** |  | 8 (19.5%) | 8 (20.0%) |
|  | *Missing, n(%)* | 0 (0.0%) | 0 (0.0%) |
| **Diabetes, n(%)** |  | 5 (12.2%) | 3 (7.9%) |
|  | *Missing, n(%)* | 0 (0.0%) | 2 (5.0%) |
| **Weakened immune system<sup>\*</sup>, n(%)** |  | 0 (0.0%) | 0 (0.0%) |
|  | *Missing, n(%)* | 0 (0.0%) | 0 (0.0%) |
| **Transplant recipient, n(%)** |  | 0 (0.0%) | 0 (0.0%) |
|  | *Missing, n(%)* | 0 (0.0%) | 0 (0.0%) |
| **Obesity, n(%)** |  | 6 (14.6%) | 2 (5.3%) |
|  | *Missing, n(%)* | 0 (0.0%) | 2 (5.0%) |
| **Mental illness, n(%)** |  | 13 (31.7%) | 13 (37.1%) |
|  | *Missing, n(%)* | 0 (0.0%) | 5 (12.5%) |
| **Hypertension, n(%)** |  | 10 (24.4%) | 5 (13.2%) |
|  | *Missing, n(%)* | 0 (0.0%) | 2 (5.0%) |
| **Any comorbidity, n(%)** |  | 34 (82.9%) | 29 (72.5%) |
|  | *Missing, n(%)* | 0 (0.0%) | 0 (0.0%) |
| <sup>*</sup> Defined as weakened immune system due to a condition participant was born with or due to |  |  |  |

disease or treatment (e.g. sickle cell, HIV, cancer, chemotherapy)

Mean symptom duration at baseline was 2.7 days (SD 1.1) in the antioxidant arm and 2.7 days (SD 1.3) in usual care. Vaccination coverage was high, with 41/41 (100.0%) in the antioxidant arm and 37/38 (97.4%) in usual care having received two or more doses. Median BMI was 27.6 (IQR 23.8–32.0) and 24.8 (IQR 23.0–30.2), respectively. Baseline symptom burden and comorbidity profiles were similar across groups, although cough was more common in the antioxidant arm at baseline.

### Change in Dietary Intake

Dietary intake data were available for all 81 participants, and 61 nutrients were analyzed. Between-group comparisons were adjusted for baseline levels **(Table 2 and Supplementary Table 1)**. No statistically significant within-group or between-group differences were observed for selenium, zinc, lycopene, or vitamin C intake from baseline to day 10. We also examined baseline supplemental intake of micronutrients and found no meaningful differences between groups. **(Table 3 and Supplementary Table 2)**.

**Table 2:** Dietary intake of selected antioxidant-related nutrients at baseline and day 10 in the antioxidant therapy and usual care groups.

| Nutrient | Arm | Baseline* | Day-10* | Mean change (95% CI) | Adjusted difference in change ($\beta$, 95% CI) |
| --- | --- | --- | --- | --- | --- |
| Selenium (mcg) | Antioxidant therapy | 85.585 ± 49.11 | 96.765 ± 45.94 | 12.500 (-2.812, 27.811) | 4.10 (-14.66, 22.86) |
| | Usual care | 83.04 ± 44.08 | 94.59 ± 40.25 | 6.454 (-8.584, 21.492) | |
| Zinc (mg) | Antioxidant therapy | 7.809 ± 4.20 | 9.18 ± 3.30 | 1.243 (-0.047, 2.532) | -0.07 (-1.86, 1.72) |
| | Usual care | 8.14 ± 4.82 | 9.37 ± 4.020 | 0.920 (-1.428, 3.268) | |
| Lycopene (mcg) | Antioxidant therapy | 2841.59 ± 3739.77 | 4913.75 ± 14881.07 | 2511.125 (-2578.233, 7600.483) | 477.67 (-6050.34, 7005.70) |
|  | Usual care | 6598.23 ± 13524.94 | 4474.78 ± 6873.17 | -3097.968 (-9486.718, 3290.783) |  |
| Vitamin C (mg) | Antioxidant therapy | 90.66 ± 142.41 | 84.24 ± 52.73 | -2.851 (-50.926, 45.224) | 0.32 (-27.73, 28.37) |
|  | Usual care | 81.92 ± 70.13 | 83.82 ± 55.79 | 2.670 (-28.304, 33.644) |  |
| *Values are presented as mean ± SD |  |  |  |  |  |

**Table 3:** Baseline supplemental intake of selected antioxidant-related nutrients.

| Nutrient | Antioxidant Therapy | Usual Care Mean |
| --- | --- | --- |
| Selenium (mcg) | 11.101 ± 18.62 | 7.781 ± 17.68 |
| Zinc (mg) | 2.038 ± 3.29 | 1.460 ± 3.28 |
| Lycopene (mg) | 9.822 ± 44.98 | 0.051 ± 0.13 |
| Vitamin C (mg) | 109.982 ± 286.605 | 185.044 ± 391.396 |

\*Values are presented as mean ± SD

Within the antioxidant arm, significant increases were observed in several other dietary variables, including total caloric intake (+202.0 kcal; *p* = 0.039), carbohydrates (+30.8 g; *p* = 0.028), folate (+53.1 mcg; *p* = 0.006), glycemic load (+17.6; *p* = 0.017), iron (+2.51 mg; *p* = 0.001), potassium (+421.7 mg; *p* = 0.005), selected B vitamins, including thiamin (*p* = 0.003), riboflavin (*p* = 0.033), and vitamin B6 (*p* = 0.013), as well as vitamin K (+33.6 mcg; *p* = 0.038). In the usual care arm, significant within-group increases were seen in total caloric intake (+236.9 kcal; *p* = 0.019), protein (+10.7 g; *p* = 0.040), and total soluble fiber (+0.62 g; *p* = 0.049). Despite these within-group changes, no adjusted between-group differences were statistically significant across any of the 61 nutrients examined **(Supplementary Table 1)**.

### Feasibility and safety outcomes

Recruitment was achieved across five Canadian provinces between September 2024 and April 2025, with 81 participants randomized. Retention was high in both groups. In the antioxidant arm, 35/41 participants (85.4%) completed 28 days of follow-up, while in the usual care arm, 33/40 (82.5%) completed 28 days of follow-up.

Adherence to the intervention was relatively high. Among participants randomized to antioxidant therapy, 37/41 (90.2%) completed the full 10-day course.

The intervention was well tolerated. Adverse events were infrequent. Seven adverse events were reported in the antioxidant arm (4 participants with at least one event), including high blood sugar, nausea, abdominal cramps, strong bad taste, heartburn and diarrhea. In the usual care arm, one adverse event (acute back pain) was reported in one participant. No serious adverse events occurred in either group. No serious adverse events were reported.

### Clinical outcomes

No participants in the antioxidant arm were hospitalized or died within 28 days (0/41, 0%), compared with 2/40 (5.0%) in the usual care arm. The Bayesian odds ratio was 0.469 (95% credible interval 0.083 to 2.38), with a probability of superiority of 0.818.

By day 14, 32/40 (80.0%) participants in the antioxidant arm had recovered compared with 23/36 (63.9%) in usual care (OR 2.128; 95% CI 0.747–4.871; probability of superiority 0.911). Complete alleviation of all symptoms by day 14 was reported by 40/40 (100.0%) in the antioxidant arm and 34/36 (94.4%) in usual care (OR 3.256; 95% CI 0.456–11.989; probability of superiority 0.841). Sustained alleviation of all symptoms occurred in 38/40 (95.0%) versus 29/36 (80.6%), respectively (OR 3.498; 95% CI 0.872–10.017; probability of superiority 0.959).

Return to usual health by day 14 was reported by 31/40 (77.5%) participants in the antioxidant arm and 22/36 (61.1%) in usual care (OR 2.241; 95% CI 0.821–5.033; probability of superiority 0.936).

Return to usual activity by day 14 occurred in 38/40 (95.0%) participants in the antioxidant arm compared with 30/36 (83.3%) in usual care (OR 3.113; 95% CI 0.762–9.022; probability of superiority 0.933) **(Table 4)**.

**Table 4:** Clinical outcomes within 28 days following randomization.

| Outcome | Antioxidant therapy<br>(N = 41) | Usual care (N<br>= 40) | Odds ratio<br>(95% CI) | Probability of<br>superiority |
| --- | --- | --- | --- | --- |
| Hospitalization or death by day<br>28 | 0 / 41 (0.0%) | 2 / 40 (5.0%) | 0.469 (0.083-2.38) | 0.818 |
| Recovery by day 14 | 32 / 40 (80.0%) | 23 / 36<br>(63.9%) | 2.128 (0.747-4.871) | 0.911 |
| Complete alleviation of all<br>symptoms by day 14 | 40 / 40 (100.0%) | 34 / 36<br>(94.4%) | 3.256 (0.456-11.989) | 0.841 |
| Sustained alleviation of all<br>symptoms | 38 / 40 (95.0%) | 29 / 36<br>(80.6%) | 3.498 (0.872-10.017) | 0.959 |
| Return to usual health by day<br>14 | 31 / 40 (77.5%) | 22 / 36<br>(61.1%) | 2.241 (0.821-5.033) | 0.936 |
| Return to usual activity by day<br>14 | 38 / 40 (95.0%) | 30 / 36<br>(83.3%) | 3.113 (0.762-9.022) | 0.933 |

Values are presented as n/N (%). Odds ratios and probability of superiority were estimated using Bayesian models. \*Bayesian odds ratio with 95% credible interval.

## Discussion

In this pilot sub-protocol embedded within the CanTreatCOVID adaptive platform trial, a 10-day course of combination antioxidant therapy was feasible to implement in a fully decentralized outpatient setting, with high adherence, good retention, and no safety concerns identified. Although no statistically significant between-group differences were observed, participants randomized to antioxidant therapy demonstrated higher proportions of recovery, symptom alleviation, and return to usual activity by day 14. The intervention was well tolerated, with no serious adverse events reported.

The biological rationale for combining selenium, zinc, vitamin C, and lycopene lies in their complementary roles in maintaining redox balance and supporting immune responses during viral infection. Acute respiratory infections are characterized by increased oxidative stress and inflammatory signaling, processes that can impair immune cell function and contribute to tissue injury. Micronutrients involved in antioxidant defense may therefore help support host resilience during viral illness.

Selenium plays a key role in endogenous antioxidant defense through its incorporation into selenoproteins such as glutathione peroxidase.^17^ These enzymes neutralize reactive oxygen species and limit oxidative damage during infection.^18^ Beyond redox buffering, selenium supports both innate and adaptive immunity, including T-cell and B-cell function and antibody production.^6^ It also modulates inflammatory pathways by promoting anti-inflammatory selenoenzymes while downregulating pro-inflammatory cytokines such as IL-6 and TNF-α.^19^ During the pandemic, ecological and observational studies suggested associations between lower selenium status and greater COVID-19 severity.^20,21^ . Although these studies cannot establish causality, they generated important hypotheses regarding selenium sufficiency and host resilience in viral infections.

Evidence from randomized trials also suggests potential benefits of selenium supplementation. A randomized, double-blind, placebo-controlled trial of 72 volunteers found that a selenium-containing formulation (100 μg/day) increased CD4+, CD3+, and CD8+ T lymphocyte levels following COVID-19 booster vaccination, without adverse events.^22^ Another multicenter randomized trial in hospitalized patients showed that supplementation with selenium (40 μg/day) as part of a nutritional formulation reduced hospital stay and time to RT-PCR negativity.^23^ Multiple mechanisms involving selenium and selenoproteins at cellular and viral levels have been implicated in influencing disease severity and recovery.^5^ However, further well-designed randomized controlled trials are needed to clarify its role in COVID treatment.

Zinc also plays a key role in antioxidant defenses and immune competence.^24^ As a component of superoxide dismutase and metallothionein, zinc contributes to neutralizing reactive oxygen species^24^, and oxidative stress during infection can mobilize intracellular zinc to buffer free radicals.^25^ Zinc has demonstrated antiviral activity *in vitro*, including inhibition of replication of several respiratory viruses.^26^ A Cochrane review of 18 trials in the common cold found that zinc reduced symptom duration by 1.03 days and was associated with reduced school absence and antibiotic use.^27^ A meta-analysis combining trial and observational data reported lower mortality with zinc supplementation (OR 0.57; 95% CI 0.43 to 0.77).^28^ More recent COVID-19 trials have reported reductions in symptom duration, ICU admission, and length of hospital stay. In a multicenter randomized trial, zinc (50 mg/day for 15 days) reduced ICU admissions and shortened hospital stay, with consistent effects across key subgroups.^29^

Vitamin C has long been studied in respiratory viral infections. Supplementation at doses exceeding 1 g/day has been associated with modest reductions in illness duration.^30^ A meta- analysis of therapeutic dosing (≥0.2 g/day) also suggested benefits for symptom duration.^31^ Observational data during the pandemic indicated that plasma vitamin C levels were often low among hospitalized patients,^32^ particularly in severe disease.^33^ In critically ill populations, vitamin C has been associated with reduced ICU stay (by 7.8% on average) and shorter duration of mechanical ventilation..^34^ Some trials have also suggested reductions in mortality among patients with sepsis^35^ and ARDS.^36^.

Lycopene, one of the most potent singlet oxygen scavengers among carotenoids, ^37^ has demonstrated anti-inflammatory and antioxidant effects in airway epithelial models. ^38^ Experimental data suggest that lycopene can reduce IL-6 expression and attenuate viral replication in cells exposed to rhinovirus^39^, a respiratory pathogen that shares inflammatory characteristics with SARS-CoV-2. Lycopene supplementation has also been shown to reduce neutrophil elastase activity in the airways.^40^ which is relevant given its role in the pathogenesis of respiratory infections including SARS-CoV2.^41^

Despite this strong biological rationale, clinical evidence supporting antioxidant supplementation for SARS-CoV-2 remains limited and inconsistent. In a systematic review and meta-analysis conducted by our group, studies evaluating antioxidants, including zinc, vitamin C, and other micronutrients, were small and heterogeneous, with most focusing on single nutrients in hospitalized patients^11^. While some studies suggested potential benefits, such as shorter symptom duration or reductions in mortality or ICU admission^42,43^, the overall certainty of the evidence was low due to small sample sizes, methodological limitations, and variability across trials.^11^ Notably, no randomized trials evaluated selenium or lycopene alone, and few assessed combination micronutrient strategies. This highlights an important gap in the literature. In contrast, our study evaluated a combination antioxidant approach administered early in infection in an outpatient population. Although not powered for definitive efficacy, the consistent directional improvements observed across symptom outcomes support the hypothesis that enhancing antioxidant defenses early in infection may impact recovery trajectories. Larger trials are needed to confirm these findings

This study has several strengths. The antioxidant arm was embedded within an established adaptive platform, enabling rapid and pragmatic evaluation without the need for a stand-alone infrastructure. The decentralized design enhanced accessibility across provinces and supported high adherence, with over 90% of participants completing the full treatment course. Baseline characteristics were well balanced. Several limitations should be considered. First, this was a pilot study with a relatively small sample size, limiting the ability to detect modest treatment effects. Second, the open-label design may introduce reporting bias in symptom-based outcomes; however, prior large pragmatic outpatient trials have not consistently found that knowledge of treatment substantially alters self-reported recovery^3,44^. While some bias cannot be excluded, the consistency across multiple symptom measures suggests it is unlikely to fully explain the findings. Third, hospitalization or death was rare, and these results should be interpreted cautiously. Finally, dietary intake was assessed using self-reported 24-hour recalls, which may be subject to recall bias and do not capture serum micronutrient levels. However, we attempted to strengthen dietary assessment by collecting repeated recalls at both baseline and day 10, including two weekdays and one weekend day at each time point, and by also collecting information on dietary supplement intake.

## Conclusions

In this pilot sub-protocol embedded within an adaptive platform trial, a 10-day course of combination antioxidant therapy was feasible to implement, well tolerated, and associated with consistent directional improvements in symptom-related outcomes among outpatients with mild to moderate SARS-CoV-2 infection. While the study was not powered to detect definitive differences in clinical endpoints, the observed trends across recovery, symptom alleviation, and return to usual activity support the biological plausibility of host-directed antioxidant strategies in early disease. Larger, adequately powered randomized trials are now needed to determine whether these signals translate into clinically meaningful benefits, and to evaluate how such interventions may complement existing outpatient therapies in the management of viral respiratory infections.

## Supporting information

Master Protocol

Antioxidant Therapy Sub-Protocol

Human Participant Research Checklist

## Declarations

### Competing interests

None to declare.

### Funding

CanTreatCOVID trial is funded by the Canadian Institutes of Health Research (CIHR) and Health Canada (Grant # FRN 183092 and PPE 190332), with the first trial therapeutic, nirmatrelvir/ritonavir (Nirmatrelvir/ritonavir), provided by the Public Health Agency of Canada. The opinions, results and conclusions reported in this article are those of the authors and are independent from any funding sources.

### Data availability statement

The data underlying this study are available from the corresponding author, BH, upon reasonable request.

### Declaration of Generative AI and AI-assisted technologies in the writing process

The author(s) declare that no generative AI or AI-assisted technologies were used in the writing of this manuscript.

### Author contributions

BH, DJ and ADP conceived of the work. BH, DJ, PD, KM, SM, AC, BRDC, MG, PJ, and PS conducted the study. NU and KS oversaw trial conduct. ML, HS and YH conducted statistical analysis of the data. DP, MP, HHMN, MM, AN, NS, GS and EB supported the dietary analysis. BH and ADP drafted the manuscript. All authors contributed to revising the manuscript for important intellectual content, gave final approval of the version to be published and agreed to be accountable for all aspects of the work.

## Acknowledgements

The authors thank our incredible patient and community partners: Brenda Andreas, Cris Carter, Jane Cooney, Gabriela Covaci, Letlotlo Gariba, Jennifer Hulme, Veronika Kiryanova, Kathy Kobow, Mike Lapenna, Mary Liu, Chris Maddison, Dorothy Nelson, Moon Ja Park, Lyric Paul, Donna Rubenstein, Dorothy Senior, Allard Schipper, Kimberly Strain, Margo Twohig, Mike Warren, John Zhan, and Alexander Zsager.

## Supplementary material

**Supplementary Table 1:** Changes in dietary intake of nutrients from baseline to day 10 in participants receiving antioxidant therapy (n=41) versus usual care (n=40).

**Supplementary Table 2:** Baseline supplemental intake of micronutrients.

## Notes

### Competing Interest Statement

The authors have declared no competing interest.

### Clinical Trial

NCT05614349

### Author Declarations

The trial was approved by Research Ethics Boards in five Canadian provinces: Unity Health Toronto Research Ethics Board – St. Michael’s Hospital (Ontario), the Conjoint Health Research Ethics Board, University of Calgary (Alberta), the University of Manitoba Biomedical Research Board – Research Ethics Bannatyne (Manitoba), the University British Columbia Children’s & Women’s Research Ethics Board (British Columbia), and the Newfoundland and Labrador Health Research Ethics Board (Newfoundland and Labrador).

