## Supplemental Table 1 and Supplemental Table 2 for "Efficacy of antioxidant therapy in mild to moderate SARS-CoV-2 infection: A pilot experimental arm of the CanTreatCOVID adaptive platform trial"

**Table S1- Changes in dietary intake of nutrients from baseline to day 10 in participants receiving antioxidant therapy (n=41) versus usual care (n=40).**

| Nutrient | Arm | Baseline mean (SD) | Day-10 mean (SD) | Mean change (95% CI) | Within Group Probability | Adjusted difference in change (β, 95% CI) |
| --- | --- | --- | --- | --- | --- | --- |
| 18:2 - Linoleic (g) | Antioxidant therapy | 8.610 (5.763) | 9.194 (4.665) | 0.486 (-1.547, 2.518) | 0.642 | -0.832 (-3.391, 1.727) |
|  | Usual care | 8.213 (5.057) | 9.985 (6.249) | 1.384 (-0.859, 3.627) | 0.238 |  |
| 18:3 - Linolenic (g) | Antioxidant therapy | 0.920 (0.774) | 0.903 (0.625) | -0.029 (-0.229, 0.172) | 0.781 | -0.247 (-0.624, 0.130) |
|  | Usual care | 0.971 (0.806) | 1.138 (1.170) | 0.223 (-0.133, 0.580) | 0.231 |  |
| 20:3 - Eicosatrienoic (g) | Antioxidant therapy | 0.005 (0.007) | 0.006 (0.009) | 0.001 (-0.002, 0.005) | 0.545 | 0.002 (-0.003, 0.006) |
|  | Usual care | 0.008 (0.011) | 0.005 (0.009) | -0.002 (-0.005, 0.002) | 0.356 |  |
| 20:4 - Arachidon (g) | Antioxidant therapy | 0.179 (0.319) | 0.171 (0.313) | -0.011 (-0.095, 0.072) | 0.79 | 0.027 (-0.111, 0.166) |
|  | Usual care | 0.243 (0.504) | 0.182 (0.297) | -0.116 (-0.327, 0.095) | 0.292 |  |
| 20:5 - EPA (g) | Antioxidant therapy | 0.073 (0.195) | 0.081 (0.190) | 0.000 (-0.063, 0.064) | 0.993 | -0.003 (-0.097, 0.091) |
|  | Usual care | 0.101 (0.264) | 0.093 (0.198) | -0.020 (-0.141, 0.102) | 0.754 |  |
| 22:5 - DPA (g) | Antioxidant therapy | 0.012 (0.029) | 0.020 (0.049) | 0.009 (-0.011, 0.028) | 0.376 | -0.005 (-0.034, 0.024) |
|  | Usual care | 0.015 (0.049) | 0.026 (0.064) | 0.019 (-0.007, 0.044) | 0.159 |  |
| 22:6 - DHA (g) | Antioxidant therapy | 0.155 (0.383) | 0.166 (0.378) | -0.000 (-0.108, 0.108) | 1 | 0.014 (-0.154, 0.181) |
|  | Usual care | 0.211 (0.542) | 0.183 (0.350) | -0.073 (-0.304, 0.159) | 0.544 |  |
| Alpha-Carotene (mcg) | Antioxidant therapy | 498.588 (703.451) | 357.702 (554.996) | -202.913 (-494.940, 89.114) | 0.182 | -166.961 (-567.850, 233.929) |
|  | Usual care | 815.651 (934.231) | 556.260 (1040.507) | -144.901 (-601.025, 311.224) | 0.539 |  |
| Beta-Carotene Equiv (mcg) | Antioxidant therapy | 3941.220 (6390.313) | 2563.596 (2493.268) | -692.862 (-1675.854, 290.131) | 0.176 | -1282.571 (-3208.604, 643.462) |
|  | Usual care | 4136.979 (4142.597) | 3881.355 (4969.873) | 348.120 (-2125.777, 2822.017) | 0.785 |  |
| Biotin (mcg) | Antioxidant therapy | 25.931 (27.158) | 23.787 (16.903) | -3.327 (-12.735, 6.080) | 0.493 | 3.170 (-4.339, 10.678) |
|  | Usual care | 16.542 (15.664) | 18.202 (11.468) | 1.811 (-1.309, 4.931) | 0.267 |  |
| Caffeine (mg) | Antioxidant therapy | 109.373 (95.533) | 124.758 (125.627) | 15.426 (-34.212, 65.064) | 0.546 | -11.639 (-80.251, 56.973) |
|  | Usual care | 108.707 (105.020) | 137.082 (160.098) | 26.310 (-23.298, 75.919) | 0.309 |  |
| Calcium (mg) | Antioxidant therapy | 659.646 (393.536) | 757.987 (370.785) | 94.221 (-13.605, 202.048) | 0.096 | 74.314 (-101.138, 249.766) |
|  | Usual care | 559.428 (286.475) | 643.345 (450.358) | 42.198 (-110.092, 194.489) | 0.592 |  |
| Calories (kcal) | Antioxidant therapy | 1392.112 (591.913) | 1607.385 (489.719) | 201.998 (17.215, 386.781)* | 0.039 | -19.270 (-270.716, 232.176) |
|  | Usual care | 1360.847 (394.709) | 1607.278 (639.279) | 236.930 (51.307, 422.554)* | 0.019 |  |
| Calories from Fat (kcal) | Antioxidant therapy | 510.211 (273.993) | 538.733 (226.185) | 29.601 (-57.509, 116.711) | 0.51 | -15.514 (-119.127, 88.100) |
|  | Usual care | 434.813 (180.204) | 530.128 (213.698) | 80.360 (-0.331, 161.051) | 0.062 |  |
| Calories from SatFat (kcal) | Antioxidant therapy | 177.714 (104.953) | 178.234 (77.570) | 2.916 (-28.924, 34.756) | 0.859 | -7.243 (-45.885, 31.399) |
|  | Usual care | 139.869 (63.421) | 171.774 (92.407) | 26.395 (-1.571, 54.361) | 0.076 |  |
| Calories from TransFat (kcal) | Antioxidant therapy | 3.909 (4.261) | 4.202 (5.162) | 0.246 (-1.618, 2.110) | 0.798 | 0.307 (-2.083, 2.698) |
|  | Usual care | 4.436 (5.656) | 3.802 (4.233) | 0.158 (-1.954, 2.271) | 0.884 |  |
| Carbohydrates (g) | Antioxidant therapy | 157.017 (71.832) | 192.790 (74.824) | 30.804 (4.553, 57.056)* | 0.028 | 0.512 (-38.867, 39.892) |
|  | Usual care | 169.668 (64.230) | 197.103 (101.499) | 27.817 (-1.627, 57.260) | 0.076 |  |
| Carotenoid RE (mcg) | Antioxidant therapy | 657.808 (1064.350) | 427.818 (416.837) | -116.135 (-280.132, 47.862) | 0.174 | -215.363 (-536.183, 105.457) |
|  | Usual care | 689.705 (690.353) | 649.043 (827.261) | 60.287 (-350.832, 471.406) | 0.776 |  |
| Choline (mg) | Antioxidant therapy | 143.955 (98.204) | 163.420 (86.130) | 29.842 (-2.537, 62.220) | 0.079 | 11.092 (-30.066, 52.249) |
|  | Usual care | 148.741 (128.092) | 155.662 (91.210) | 13.528 (-25.436, 52.491) | 0.502 |  |
| Fat (g) | Antioxidant therapy | 56.804 (30.519) | 59.971 (25.226) | 3.298 (-6.407, 13.003) | 0.51 | -1.710 (-13.253, 9.832) |
|  | Usual care | 48.360 (20.021) | 58.974 (23.761) | 8.957 (-0.010, 17.924) | 0.062 |  |
| Folate (mcg) | Antioxidant therapy | 250.340 (162.470) | 284.203 (91.099) | 53.135 (17.274, 88.996)** | 0.006 | -20.248 (-92.540, 52.044) |
|  | Usual care | 294.348 (148.053) | 319.206 (180.194) | 15.279 (-69.161, 99.719) | 0.726 |  |
| Folate, DFE (mcg DFE) | Antioxidant therapy | 288.280 (180.292) | 338.069 (124.318) | 70.954 (26.226, 115.683)** | 0.004 | -10.929 (-99.214, 77.356) |
|  | Usual care | 349.373 (199.133) | 361.978 (208.312) | -1.598 (-114.602, 111.405) | 0.978 |  |
| Folate, food (mcg) | Antioxidant therapy | 190.285 (150.910) | 201.439 (70.855) | 25.806 (-2.778, 54.390) | 0.086 | -20.890 (-81.429, 39.650) |
|  | Usual care | 210.286 (122.139) | 235.802 (169.031) | 26.405 (-39.955, 92.765) | 0.443 |  |
| Folic Acid (mcg) | Antioxidant therapy | 52.161 (50.782) | 75.718 (63.685) | 28.159 (11.139, 45.178)** | 0.003 | 9.464 (-25.613, 44.541) |
|  | Usual care | 76.088 (106.072) | 71.504 (70.971) | -16.224 (-71.329, 38.880) | 0.569 |  |
| Glycemic Load | Antioxidant therapy | 76.605 (37.084) | 96.529 (40.463) | 17.581 (3.795, 31.368)* | 0.017 | 0.525 (-21.385, 22.434) |
|  | Usual care | 86.152 (36.004) | 101.277 (56.417) | 14.187 (-3.407, 31.780) | 0.127 |  |
| Iodine (mcg) | Antioxidant therapy | 63.773 (42.216) | 68.029 (51.660) | 1.215 (-10.704, 13.133) | 0.843 | -0.447 (-19.980, 19.086) |
|  | Usual care | 48.887 (36.798) | 54.889 (40.388) | 5.395 (-9.540, 20.329) | 0.486 |  |
| Iron (mg) | Antioxidant therapy | 9.251 (4.218) | 11.673 (3.732) | 2.511 (1.200, 3.821)** | 0.001 | 1.038 (-1.138, 3.215) |
|  | Usual care | 10.995 (5.452) | 11.087 (4.855) | -0.155 (-2.960, 2.650) | 0.914 |  |
| Lutein & Zeaxanthin (mcg) | Antioxidant therapy | 1367.276 (2016.252) | 1306.465 (1507.139) | 57.231 (-714.962, 829.423) | 0.885 | -464.061 (-1593.611, 665.489) |
|  | Usual care | 1642.356 (1650.258) | 1793.603 (2861.703) | 306.528 (-875.173, 1488.228) | 0.616 |  |
| Lycopene (mcg) | Antioxidant therapy | 2841.595 (3739.777) | 4913.752 (14881.070) | 2511.125 (-2578.233, 7600.483) | 0.34 | 477.678 (-6050.347, 7005.702) |
|  | Usual care | 6598.232 (13524.942) | 4474.783 (6873.178) | -3097.968 (-9486.718, 3290.783) | 0.351 |  |
| Magnesium (mg) | Antioxidant therapy | 257.857 (146.973) | 295.816 (115.041) | 28.712 (-13.029, 70.454) | 0.186 | -17.385 (-89.832, 55.062) |
|  | Usual care | 259.207 (115.204) | 306.247 (203.001) | 51.115 (-16.104, 118.334) | 0.149 |  |
| Mono Fat (g) | Antioxidant therapy | 21.366 (13.448) | 23.224 (11.641) | 1.858 (-2.610, 6.325) | 0.421 | 0.660 (-4.397, 5.716) |
|  | Usual care | 17.484 (7.982) | 21.335 (8.566) | 3.487 (-0.385, 7.358) | 0.09 |  |
| MyPlate - Dairy (c) | Antioxidant therapy | 1.369 (1.101) | 1.483 (1.159) | 0.094 (-0.278, 0.467) | 0.623 | 0.096 (-0.396, 0.588) |
|  | Usual care | 0.820 (0.686) | 1.099 (1.082) | 0.144 (-0.123, 0.411) | 0.301 |  |
| MyPlate - Fruit (c) | Antioxidant therapy | 0.947 (1.005) | 0.889 (0.826) | -0.150 (-0.427, 0.127) | 0.296 | -0.141 (-0.550, 0.269) |
|  | Usual care | 0.812 (0.887) | 0.917 (1.123) | 0.073 (-0.284, 0.431) | 0.691 |  |
| MyPlate - Grain Total (oz-eq) | Antioxidant therapy | 4.744 (2.857) | 5.921 (2.965) | 1.134 (0.129, 2.138)* | 0.034 | 0.475 (-1.128, 2.078) |
|  | Usual care | 5.838 (3.524) | 6.153 (4.152) | 0.084 (-1.340, 1.508) | 0.909 |  |
| MyPlate - Protein Total (oz-eq) | Antioxidant therapy | 5.090 (3.648) | 5.920 (3.422) | 0.771 (-0.730, 2.272) | 0.321 | -0.794 (-2.474, 0.886) |
|  | Usual care | 5.485 (3.013) | 6.756 (3.073) | 1.348 (-0.170, 2.866) | 0.094 |  |
| MyPlate - Vegetable Total (c) | Antioxidant therapy | 1.069 (1.305) | 1.388 (0.856) | 0.481 (0.173, 0.790)** | 0.004 | -0.322 (-0.933, 0.289) |
|  | Usual care | 1.355 (1.259) | 1.893 (1.621) | 0.508 (-0.175, 1.192) | 0.157 |  |
| Net Carbs (g) | Antioxidant therapy | 139.440 (64.771) | 173.312 (70.085) | 29.363 (5.275, 53.450)* | 0.022 | 1.399 (-34.904, 37.702) |
|  | Usual care | 151.431 (60.050) | 177.480 (93.217) | 25.191 (-2.057, 52.439) | 0.082 |  |
| Omega 3 Fatty Acid (g) | Antioxidant therapy | 1.274 (1.186) | 1.291 (1.103) | -0.026 (-0.336, 0.283) | 0.868 | -0.182 (-0.638, 0.275) |
|  | Usual care | 1.309 (1.253) | 1.460 (1.320) | 0.162 (-0.240, 0.565) | 0.436 |  |
| Omega 6 Fatty Acid (g) | Antioxidant therapy | 9.304 (6.206) | 9.775 (4.935) | 0.322 (-1.890, 2.535) | 0.777 | -1.095 (-3.933, 1.744) |
|  | Usual care | 8.612 (5.256) | 10.722 (7.037) | 1.671 (-0.839, 4.180) | 0.204 |  |
| Pantothenic Acid (mg) | Antioxidant therapy | 4.044 (2.078) | 4.453 (1.556) | 0.409 (-0.168, 0.987) | 0.173 | -0.489 (-1.484, 0.507) |
|  | Usual care | 4.332 (3.104) | 5.102 (2.614) | 0.517 (-0.861, 1.895) | 0.469 |  |
| Phosphorus (mg) | Antioxidant therapy | 1072.753 (563.174) | 1212.887 (437.136) | 119.600 (-39.084, 278.284) | 0.149 | 4.675 (-209.359, 218.709) |
|  | Usual care | 1027.295 (358.129) | 1180.067 (515.135) | 142.333 (-27.835, 312.502) | 0.114 |  |
| Poly Fat (g) | Antioxidant therapy | 10.908 (6.703) | 11.345 (5.805) | 0.237 (-2.038, 2.513) | 0.839 | -1.653 (-4.775, 1.470) |
|  | Usual care | 10.502 (6.013) | 12.923 (7.888) | 1.980 (-0.761, 4.721) | 0.169 |  |
| Potassium (mg) | Antioxidant therapy | 2078.438 (1154.250) | 2501.310 (856.417) | 421.653 (144.401, 698.905)** | 0.005 | 73.994 (-366.497, 514.486) |
|  | Usual care | 2167.309 (751.562) | 2464.748 (1113.353) | 315.539 (-90.315, 721.393) | 0.14 |  |
| Protein (g) | Antioxidant therapy | 65.131 (35.182) | 74.339 (23.234) | 9.113 (-1.088, 19.315) | 0.089 | -0.642 (-12.383, 11.099) |
|  | Usual care | 63.207 (21.307) | 74.403 (27.969) | 10.660 (1.026, 20.294)* | 0.04 |  |
| Saturated Fat (g) | Antioxidant therapy | 19.746 (11.661) | 19.804 (8.618) | 0.324 (-3.214, 3.862) | 0.859 | -0.804 (-5.098, 3.489) |
|  | Usual care | 15.540 (7.046) | 19.085 (10.268) | 2.933 (-0.175, 6.040) | 0.076 |  |
| Selenium (mcg) | Antioxidant therapy | 85.585 (49.109) | 96.765 (45.938) | 12.500 (-2.812, 27.811) | 0.119 | 4.100 (-14.663, 22.864) |
|  | Usual care | 83.040 (44.084) | 94.588 (40.248) | 6.454 (-8.584, 21.492) | 0.408 |  |
| Sodium (mg) | Antioxidant therapy | 1989.030 (1017.109) | 2385.034 (1168.159) | 418.858 (4.688, 833.029) | 0.055 | 291.483 (-241.075, 824.040) |
|  | Usual care | 2314.075 (1205.493) | 2256.793 (923.707) | -202.684 (-688.318, 282.949) | 0.421 |  |
| Total Dietary Fiber (g) | Antioxidant therapy | 17.574 (10.570) | 19.474 (7.662) | 1.441 (-1.675, 4.556) | 0.371 | -0.742 (-5.031, 3.546) |
|  | Usual care | 18.238 (8.626) | 19.621 (11.912) | 2.623 (-0.568, 5.815) | 0.12 |  |
| Total Soluble Fiber (g) | Antioxidant therapy | 2.347 (2.045) | 2.609 (1.252) | 0.231 (-0.512, 0.975) | 0.546 | -0.090 (-0.829, 0.649) |
|  | Usual care | 2.273 (2.142) | 2.597 (1.783) | 0.617 (0.034, 1.199)* | 0.049 |  |
| Trans Fatty Acid (g) | Antioxidant therapy | 0.435 (0.474) | 0.467 (0.574) | 0.028 (-0.179, 0.235) | 0.794 | 0.035 (-0.231, 0.301) |
|  | Usual care | 0.494 (0.628) | 0.422 (0.471) | 0.017 (-0.218, 0.252) | 0.889 |  |
| Vitamin A - IU (IU) | Antioxidant therapy | 7477.300 (10741.911) | 5339.330 (4416.194) | -1000.980 (-2723.043, 721.083) | 0.262 | -2131.646 (-5316.787, 1053.495) |
|  | Usual care | 7571.550 (6745.380) | 7515.915 (8037.627) | 874.231 (-3094.550, 4843.012) | 0.67 |  |
| Vitamin B1 - Thiamin (mg) | Antioxidant therapy | 1.040 (0.543) | 1.301 (0.456) | 0.266 (0.100, 0.431)** | 0.003 | 0.015 (-0.368, 0.398) |
|  | Usual care | 1.178 (0.540) | 1.357 (1.041) | 0.143 (-0.270, 0.555) | 0.504 |  |
| Vitamin B12 (mcg) | Antioxidant therapy | 2.449 (2.071) | 3.182 (2.319) | 0.678 (-0.122, 1.478) | 0.106 | 0.131 (-0.879, 1.141) |
|  | Usual care | 2.504 (2.517) | 3.151 (1.616) | 0.280 (-0.835, 1.395) | 0.627 |  |
| Vitamin B2 - Riboflavin (mg) | Antioxidant therapy | 1.414 (0.648) | 1.650 (0.606) | 0.236 (0.028, 0.444)* | 0.033 | 0.076 (-0.227, 0.378) |
|  | Usual care | 1.409 (0.653) | 1.605 (0.703) | 0.121 (-0.171, 0.412) | 0.424 |  |
| Vitamin B3 - Niacin Equiv (mg) | Antioxidant therapy | 26.873 (16.274) | 31.115 (10.845) | 4.565 (-1.160, 10.290) | 0.127 | -1.293 (-7.655, 5.068) |
|  | Usual care | 27.182 (11.357) | 32.990 (14.753) | 3.870 (-1.847, 9.586) | 0.197 |  |
| Vitamin B6 (mg) | Antioxidant therapy | 1.224 (0.799) | 1.504 (0.555) | 0.304 (0.075, 0.532)* | 0.013 | -0.007 (-0.370, 0.356) |
|  | Usual care | 1.390 (0.767) | 1.615 (0.951) | 0.141 (-0.239, 0.521) | 0.473 |  |
| Vitamin C (mg) | Antioxidant therapy | 90.661 (142.413) | 84.246 (52.736) | -2.851 (-50.926, 45.224) | 0.908 | 0.319 (-27.733, 28.371) |
|  | Usual care | 81.920 (70.135) | 83.826 (55.790) | 2.670 (-28.304, 33.644) | 0.867 |  |
| Vitamin D - IU (IU) | Antioxidant therapy | 105.110 (170.308) | 137.910 (185.132) | 24.028 (-55.326, 103.382) | 0.557 | 34.836 (-48.199, 117.871) |
|  | Usual care | 69.527 (70.419) | 97.246 (114.912) | 24.400 (-23.015, 71.814) | 0.323 |  |
| Vitamin E - IU (IU) | Antioxidant therapy | 9.822 (8.373) | 9.931 (7.017) | 0.076 (-2.306, 2.458) | 0.95 | 0.547 (-2.647, 3.741) |
|  | Usual care | 9.497 (8.158) | 9.534 (7.490) | -0.665 (-3.962, 2.631) | 0.696 |  |
| Vitamin K (mcg) | Antioxidant therapy | 75.562 (137.992) | 91.758 (86.242) | 33.609 (2.996, 64.223)* | 0.038 | -1.588 (-56.237, 53.061) |
|  | Usual care | 91.607 (90.082) | 106.791 (133.976) | 18.971 (-31.647, 69.589) | 0.469 |  |
| Zinc (mg) | Antioxidant therapy | 7.809 (4.199) | 9.179 (3.306) | 1.243 (-0.047, 2.532) | 0.067 | -0.072 (-1.867, 1.723) |
|  | Usual care | 8.148 (4.825) | 9.372 (4.020) | 0.920 (-1.428, 3.268) | 0.45 |  |

**Table S2 – Baseline supplemental intake of micronutrients**

| **Nutrient** | **Antioxidant Therapy Mean (SD)** | **Usual Care Mean (SD)** |
| --- | --- | --- |
| Vitamin A (IU) | 281.951 (516.472) | 169.464 (458.987) |
| Vitamin E (IU) | 9.120 (14.496) | 44.273 (193.022) |
| Beta-Carotene (IU) | 399.964 (742.099) | 285.558 (712.061) |
| Folate (mg) | 0.184 (0.341) | 0.180 (0.322) |
| Calcium (mg) | 95.526 (197.310) | 154.287 (384.333) |
| Iron (mg) | 9.233 (30.357) | 4.012 (12.549) |
| Vitamin B1 (mg) | 12.627 (30.818) | 13.475 (36.547) |
| Vitamin B2 (mg) | 11.498 (29.908) | 9.488 (20.639) |
| Vitamin B5 (mg) | 10.169 (28.548) | 4.487 (16.645) |
| Vitamin B6 (mg) | 11.724 (29.880) | 6.747 (17.589) |
| Vitamin B12 (mcg) | 108.606 (310.507) | 100.899 (253.701) |
| Vitamin D (IU) | 181.405 (295.503) | 139.923 (314.333) |
| Vitamin K1 (mcg) | 5.820 (12.169) | 3.195 (7.840) |
| Biotin (mcg) | 21.866 (37.755) | 20.946 (38.935) |
| Choline (mg) | 10.479 (29.230) | 4.212 (16.625) |
| Chromium (mcg) | 16.941 (29.587) | 12.672 (31.075) |
| Copper (mcg) | 145.971 (285.558) | 106.906 (263.794) |
| Iodine (mcg) | 33.518 (52.137) | 24.629 (54.126) |
| Lutein (mg) | 13.526 (60.174) | 0.074 (0.185) |
| Magnesium (mg) | 20.248 (34.206) | 13.989 (33.215) |
| Manganese (mg) | 0.649 (1.232) | 0.414 (0.976) |
| Molybdenum (mcg) | 7.110 (14.857) | 6.800 (16.433) |
| Niacinamide (mg) | 13.417 (30.192) | 11.622 (23.656) |
| Pathothenic Acid (mg) | 12.109 (29.580) | 6.211 (16.998) |
| Phosphorus (mg) | 0.265 (1.695) | 0.000 (0.000) |
| Potassium (mg) | 1.428 (8.917) | 0.571 (3.612) |
