## Supplementary material for "Efficacy of antioxidant therapy in mild to moderate SARS-CoV-2 infection: A pilot experimental arm of the CanTreatCOVID adaptive platform trial": Master Protocol

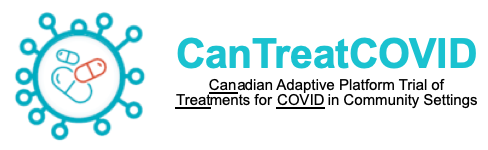

**Can**adian Adaptive Platform Trial of **Treat**ments for **COVID** in Community Settings

**Principal Investigator:** Andrew D. Pinto MD CCFP FRCPC

**Contact Details:** Upstream Lab, MAP/Centre for Urban Health Solutions, Li Ka Shing Knowledge Institute, Unity Health Toronto, 30 Bond Street, Toronto, Ontario, Canada M5B1W8,, 416-864-6060 x76148

**Study Sponsor:** Unity Health Toronto

**Contact Details:**

**Study Funder:** Canadian Institutes for Health Research (CIHR)

**Contact Details:**

**Date:** March 26, 2025

**Protocol Version:** 6.0

**Clinical Trials.gov identifier:** NCT05614349

**SPONSOR SIGNATURE PAGE**

Title: Canadian Adaptive Platform Trial of Treatments for COVID in Community Settings (CanTreatCOVID)

CanTreatCOVID is an open-label, individually randomized, multi-centre, national trial. CanTreatCOVID aims to establish an adaptive platform trial aimed at evaluating the clinical- and cost-effectiveness, practical challenges, and outcomes of therapeutics for SARS-CoV-2 for non-hospitalized patients in Canada. Participants will be randomized to receive usual care (i.e., supportive care and symptom relief) or a study therapeutic, which will be determined by the Canadian COVID-19 Out-Patient Therapeutics Committee. The primary outcomes being evaluated is all-cause hospitalization and/or death at 28 days.

I agree to the terms and conditions relating to this study as defined in this protocol. I will conduct this study as outlined and will make a reasonable effort to complete the study within the time designated.

I agree to conduct this study in accordance with the declaration of Helsinki and its amendments, the Tri-Council Policy Statement: Ethical Conduct for Research Involving Humans (TCPS-2), International Council for Harmonisation (ICH) Good Clinical Practice Guidelines (GCP) and applicable regulations and laws. I will obtain the approval of a Research Ethics Board for this protocol prior to its implementation.

I have read this protocol and agree that it contains all the necessary details for carrying out this study. I will conduct the study as outlined herein and will complete this study within the time designated.

I will provide copies of the protocol and all pertinent information to all individuals responsible to me who assist in the conduct of this study. I will discuss this material with them to ensure that they are fully informed and trained regarding the study drug(s), the conduct, and the obligations of confidentiality as per the Canadian Privacy Act, The Personal Information Protection and Electronic Documents Act (“PIPEDA”) and the relevant HealthCare Privacy Legislations.

I confirm that I will conduct this clinical trial in compliance with the International Council for Harmonisation Good Clinical Practice Guideline (ICH-GCP E6), the Tri-Council Policy Statement: Ethical Conduct for Research Involving Humans (TCPS-2), applicable Health Canada regulations, the Declaration of Helsinki, the Protocol as approved, and all applicable local and study specific standard operating procedures (SOPs).

**Sponsor Name:** Andrew D. Pinto

**Sponsor Signature:**
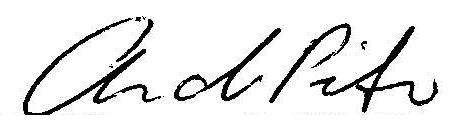

**Date:** August 23, 2023

**SITE INVESTIGATOR SIGNATURE PAGE**

Title: Canadian Adaptive Platform Trial of Treatments for COVID in Community Settings (CanTreatCOVID)

CanTreatCOVID is an open-label, individually randomized, multi-centre, national trial. CanTreatCOVID aims to establish an adaptive platform trial aimed at evaluating the clinical- and cost-effectiveness, practical challenges, and outcomes of therapeutics for SARS-CoV-2 for non-hospitalized patients in Canada. Participants will be randomized to receive usual care (i.e. supportive care and symptom relief) or a study therapeutic, which will be determined by the Canadian COVID-19 Out-Patient Therapeutics Committee. The primary outcome being evaluated is all-cause hospitalization and/or death at 28 days.

I agree to the terms and conditions relating to this study as defined in this protocol. I will conduct this study as outlined and will make a reasonable effort to complete the study within the time designated.

I agree to conduct this study in accordance with the declaration of Helsinki and its amendments, the Tri-Council Policy Statement: Ethical Conduct for Research Involving Humans (TCPS-2), International Council for Harmonisation (ICH) Good Clinical Practice Guidelines (GCP) and applicable regulations and laws. I will obtain the approval of a Research Ethics Board for this protocol prior to its implementation.

I have read this protocol and agree that it contains all the necessary details for carrying out this study. I will conduct the study as outlined herein and will complete this study within the time designated.

I will provide copies of the protocol and all pertinent information to all individuals responsible to me who assist in the conduct of this study. I will discuss this material with them to ensure that they are fully informed and trained regarding the study drug(s), the conduct, and the obligations of confidentiality as per the Canadian Privacy Act, The Personal Information Protection and Electronic Documents Act (“PIPEDA”) and the relevant HealthCare Privacy Legislations.

I confirm that I will conduct this clinical trial in compliance with the International Council for Harmonisation Good Clinical Practice Guideline (ICH-GCP E6), the Tri-Council Policy Statement: Ethical Conduct for Research Involving Humans (TCPS-2), applicable Health Canada regulations, the Declaration of Helsinki, the Protocol as approved, and all applicable local and study specific standard operating procedures (SOPs).

| **Site Investigator Name** | **Signature** | **Date** |
| --- | --- | --- |

### 1. OVERVIEW

| **Title** | **Can**adian Adaptive Platform Trial of **Treat**ments for **COVID** in Community Settings (CanTreatCOVID) |
| --- | --- |
| **Study design** | Open-label, individually randomized, adaptive platform trial |
| **Study duration** | Perpetual |
| **Participant duration** | 24 months |
| **Objectives** | 1) Establish an adaptive platform trial aimed at evaluating the effectiveness (including comparative clinical and cost-effectiveness), practical challenges, and outcomes of therapeutics for SARS-CoV-2for non-hospitalized patients in Canada, engaging a variety of healthcare settings.  2) Generate evidence on treatment effectiveness and outreach to communities made vulnerable by social and economic policies, particularly those historically excluded from research.  3) Provide rapid evidence to inform clinical and health system management and public health leaders, decision-makers, and planners within Canada and internationally. |
| **Expected number of participants** | This adaptive platform trial incorporates Bayesian adaptive design principles, where the ultimate number of arms can change and the ultimate number of participants in each arm is not established using a fixed, *a priori* calculated sample size. |
| **Inclusion and exclusion criteria** | Inclusion: Age 50 years and older or 18-49 with 1 or more chronic high-risk medical conditions or immunosuppression within 5 days of onset of SARS-CoV-2 symptoms.  Exclusion: Currently admitted to hospital or in an emergency department (ED) for more than 24 hours, previously randomized to CanTreatCOVID, currently participating in a clinical trial of a therapeutic agent for acute SARS-CoV-2 infection that is not/suspected not compatible with the study therapeutics, already taking a study therapeutic or contraindication to a study therapeutic, or inability for participant or caregiver to provide informed consent. |
| **Study interventions** | Usual Care  *Therapeutics for SARS-CoV-2 in out-patient settings will be added, to be determined by the Canadian COVID-19 Out-Patient Therapeutics Committee* |
| **Methodology** | **Outcome (primary):** All cause hospitalization or death at 28 days **Outcome (secondary: all cause emergency department visits,** time to recovery; symptom severity; incidence of post-acute sequelae of SARS-CoV-2; quality of life; costs and cost/QALY |

### 2. ABSTRACT

While public health measures and vaccines have reduced the impact of SARS-CoV-2 on hospitalization and death, most scientists predict this virus will become endemic and new variants will continue to emerge. Effective and affordable therapeutics for SARS-CoV-2 that can be easily used in community settings are needed to accelerate recovery, prevent hospitalizations and deaths, and to minimize the development of post-acute sequelae of SARS-CoV-2 (“long COVID”). Most randomized controlled trials (RCT) of therapeutics to date have included participants who have not been vaccinated and who did not have previous infections. The **Can**adian Adaptive Platform Trial of **Treat**ments for **COVID** in Community Settings (CanTreatCOVID) will evaluate the clinical effectiveness and cost-effectiveness of therapeutics for SARS-CoV-2 in non-hospitalized patients. Adaptive platform trials (APTs) are designed to compare multiple therapies in an efficient manner and allow us to respond to the dynamic nature of the COVID-19 pandemic. Therapeutics to be evaluated will be identified through a transparent Canadian COVID-19 Out-Patient Therapeutics Committee. The primary outcome is all-cause hospitalization and/or death at 28 days, and key secondary outcomes include time to recovery, symptom severity, incidence of post-acute sequelae of SARS-CoV-2, quality of life, and cost-effectiveness of each therapeutic. CanTreatCOVID uses numerous approaches to recruit participants to the study, including a multi-faceted public communication strategy and outreach through primary care, out-patient clinics, and EDs.

### 3. BACKGROUND AND RATIONALE

As the world enters the third year of the COVID-19 pandemic, over 6.4 million people globally have died of SARS-CoV-2.^1^ While vaccines have reduced the proportion of patients requiring hospitalization and critical care and thus prevented health systems from being overwhelmed, SARS-CoV-2 will likely become endemic.^2^ New variants will emerge and reduced vaccine effectiveness against future strains remains a concern.^3^ While monoclonal antibody treatments have been effective, several (e.g. bamlanivimab, casirivimab/imdevimab, and sotrovimab) have already proven vulnerable to antiviral resistance.^4,5^ As with certain antivirals (e.g. remdesivir), they require IV infusion, are expensive, and are usually administered in specialized centres for outpatient treatment in those at highest risk.^6^ **Effective, safe, convenient, affordable, and evidence-based therapeutics that can be used in communities with high vaccination rates to limit the severity of SARS-CoV-2 infection, reduce hospitalizations, and reduce short- and long-term symptoms remain urgently needed**.^7^

Studies to date have evaluated numerous potential agents in community settings for mild to moderate infections, but current Canadian^8,9^, American,^10^ and international guidelines^11,12^ identify only nirmatrelvir/ritonavir (Paxlovid™) as a recommended therapeutic.^12^ Fluvoxamine has moderate evidence,^13^ inhaled steroids have weak evidence but may be considered,^12,14^ and molnupiravir is currently under Health Canada review.^15^ Nirmatrelvir/ritonavir is a promising treatment, however the key RCT that evaluated this treatment, the Evaluation of Protease Inhibition for COVID-19 in High-Risk Patients (EPIC-HR) study, was in unvaccinated patients, and predated the most recent variants of SARS-CoV-2.^16^ Subsequent data released from the EPIC-SR study was not able to demonstrate efficacy in the primary outcome of time to recovery, nor in the key secondary outcome of hospitalization or death at 28 days. The sub-group analysis for 721 vaccinated adults with at least one risk factor for progression to severe COVID-19 demonstrated a non-significant relative risk reduction (RR 0.43; 95%CI 0.11-1.64).^17^ A recent systematic review and network meta-analysis of antivirals for non-severe SARS-CoV-2 infection concluded that it was essential to evaluate the effectiveness of nirmatrelvir/ritonavir and other new agents in the vaccinated population and against more recent variants.^18^

Three major problems are currently faced by clinicians, provincial decision makers, public health leaders, and patients:^19^

i. Almost all published trials have included only unvaccinated patients. **It is unclear whether and to what extent existing therapeutics are effective in partially or fully vaccinated patients, or among those with prior infection.**
ii. **Therapeutics have not been compared to one another**, and the comparative effectiveness, safety and cost-effectiveness/cost-utility has not been established.^20^
iii. Currently, **no therapeutic has been evaluated specifically for its potential in reducing the likelihood of post-acute sequelae of SARS-CoV-2**.

***Why is an APT for COVID therapeutics in community settings needed now?***

First, vaccines against SARS-CoV-2 have reduced infection severity and significantly fewer patients are presenting to emergency rooms and hospitals.^21^ Patients with mild to moderate symptoms – including those at high risk of deterioration – present to primary care and other community settings.^22^ This APT's recruitment strategies reflect settings where patients present early in the course of infection and where decisions about prescriptions are likely to be made, enhancing generalizability. Clinicians and patients need to know which therapeutics are the most effective. As approximately 85% of Canadians have access to a regular family physician or source of primary care,^23^ it is likely that the prescribing of COVID therapeutics for a significant proportion of our target population will occur in that context. Effective therapeutics could significantly reduce the burden on EDs, hospitals, and intensive care units (ICUs) if case numbers rise and/or vaccine efficacy against severe disease wanes. Second, Canadian decision makers currently have limited evidence to inform decisions on procurement^24,25^ and comparative efficacy trials are needed. Third, a nimble and durable trial structure is needed, such that Canadians do not have to rely on trials carried out in other countries and can generate their own data in real time.

**APTs are an ideal design to compare multiple therapies and to allow for new therapeutics to be added to the trial as they emerge**. APTs study multiple interventions in a single disease/condition in a perpetual manner, with new research questions focused on new interventions allowed to enter or leave the platform over time based on a decision algorithm. APTs use a master protocol to establish standard operating procedures and create a common clinical trial evaluation for all interventions, including interim analyses to determine whether enrollment to interventions can be stopped early for statistical efficiencies (sequential designs). During the COVID-19 pandemic, APTs have been crucial in identifying what does and does not work to treat SARS-CoV-2 for hospitalized patients.^26^ For example, the RECOVERY Trial helped identify that dexamethasone significantly reduced in mortality among ventilated patients.^27^ REMAP-CAP identified the important role of intravenous steroids in ICU patients.^28^ Later in collaboration with other APTs, ATTACC, REMAP-CAP, and ACTIV-IV investigators demonstrated therapeutic-dose heparin reduced progression to ICU level organ support and death in non-critically ill hospitalized patients with COVID-19.^29,30^ Among patients seen in the community, the PRINCIPLE trial in the UK demonstrated the efficacy of inhaled budesonide.^31^

### 4. STUDY OBJECTIVES AND RESEARCH QUESTIONS

1) Establish an APT aimed at evaluating the effectiveness (including comparative clinical and cost-effectiveness), practical challenges, and outcomes of therapeutics for SARS-CoV-2 to non-hospitalized patients in Canada, engaging a variety of healthcare settings.
2) Generate evidence on treatment effectiveness and outreach to communities made vulnerable by social and economic policies, particularly those historically excluded from research.
3) Provide rapid evidence to inform clinical and health system management and public health leaders, decision-makers, and planners within Canada and internationally.

***Central research questions:*** *What is the comparative effectiveness of SARS-CoV-2 therapeutics for non-hospitalized patients? What is the comparative effectiveness in reducing post-acute sequelae of SARS-CoV-2? Are the tested therapeutics cost-effective?* *How does clinical- and cost-effectiveness differ by underlying patient risk and across diverse populations?*

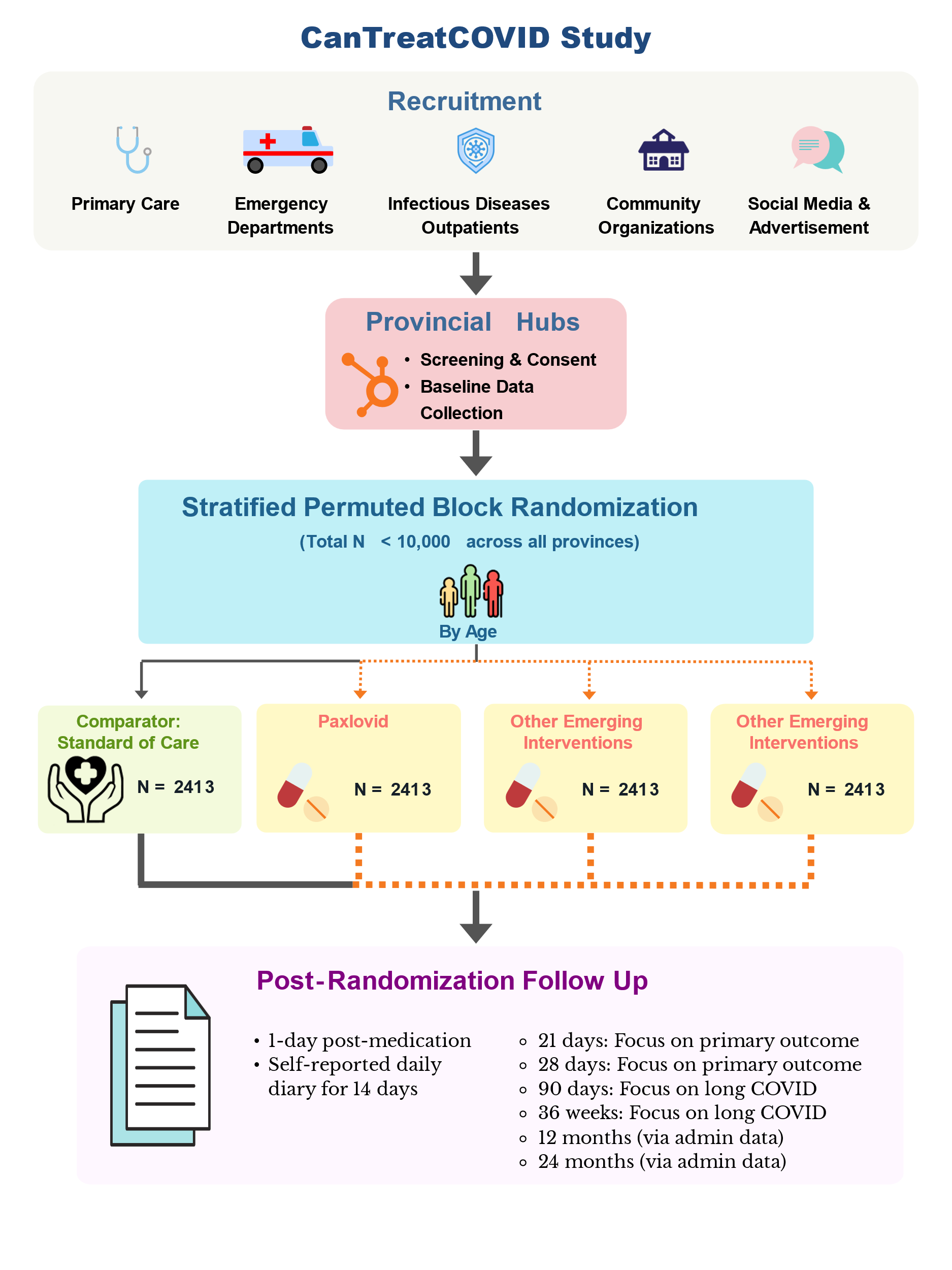
***Figure 1: CanTreatCOVID study diagram.***

Excluded for follow ups past April 28th 2025.

### 5. STUDY DESIGN AND METHODOLOGY

#### 5.1 **Study design**

CanTreatCOVID is an open-label, individually randomized, adaptive platform trial enrolling high-risk patients (50+ years old or 18-49 with 1+ chronic medical conditions and/or immunosuppression) within 5 days of symptom onset to evaluate therapeutics for SARS-CoV-2 infection against usual care and among each other. Therapeutics for SARS-CoV-2 in out-patient settings to be evaluated will be determined by the Canadian COVID-19 Out-Patient Therapeutics Committee. The design will allow adaptations to the trial based on primary outcome data, including removal of intervention arms based on declarations of success or futility at an interim analysis, integration of results from international trials, and adding intervention arms.

**Population:**	Patient population deemed at moderate to high risk of progression to severe disease by current Canadian data, currently: older adults (50+) years old or 18-49 with 1+ chronic medical condition and/or who are immunosuppressed with positive SARS-CoV-2 test (polymerase chain reaction (PCR) or rapid antigen test (RAT)), within 5 days of symptom onset

**Intervention:**	*Therapeutics for SARS-CoV-2 in out-patient settings to be evaluated will be determined by the Canadian COVID-19 Out-Patient Therapeutics Committee*

**Control:**	Usual care

**Outcome (primary):**	All cause hospitalization or death at 28 days

**Outcome (secondary):** All cause ED visits, Time to recovery; Symptom severity; incidence of post-acute sequelae of SARS-CoV-2; quality of life; costs and cost/QALY

#### 5.2 **Endpoints and outcomes**

***Primary outcome:*** All cause **Hospitalization or death at 28 days** from randomization, captured during participant follow up and corroborated with administrative data. Based on a robust understanding of SARS-CoV-2 infection, it is likely that severe outcomes would occur within 28 days of symptom onset^32,33^, and this outcome has been used in several key studies of SARS-CoV-2 treatments in community settings.^31,34–36^

***Secondary outcomes:***

- All cause emergency-department visit
- **Time to recovery**, using the questions: “Do you feel recovered today? (i.e. symptoms associated with illness are no longer a problem), used in PRINCIPLE and PANORAMIC^31,36^; and Flu Pro Plus questions about returning to usual health and activities, which can be used to determine time to recovery.^37^
- **Symptom severity**, using the questions: “*How well are you feeling today? Please rate how you are feeling now using a scale of 1 – 4, where 1 is no symptoms, and 4 is very severe symptoms”* and by rating symptoms, if present, as *“No problem, mild problem, moderate problem, or major problem.”*
- **Health service use, treatment costs, cost/QALY**
- Quality of life, using **EQ-5D-5L** and analyzed using Canadian reference values. Participants will be followed up until April 28^th^, 2025.
- **Post-acute sequelae of SARS-CoV-2**, using the World Health Organization clinical case definition^38,39^ and the recently validated Symptom Burden Questionnaire for Long COVID^40.^ Participants will be followed up until April 28^th^, 2025.

**Early discontinuation and severe adverse events.**

#### 5.3 **Measures to minimize bias, randomization and blinding**

In this open-label adaptive platform trial, the participant and recruiting clinicians will know which intervention is being used, but outcome adjudication, analysis, and the investigators will remain blinded.

As part of the adaptive design, the interim results will be monitored by the independent statistical center and independent Data Safety and Monitoring Committee (DSMC) [Section 19]. The DSMC will assess whether the randomized comparisons in the study have provided evidence that is strong enough on the primary outcome of interest, once finalized. New arms can be added as evidence emerges that other candidate therapeutics are deemed as worthy of evaluation by the central prioritization committee. Others including the Steering Committee will not have access to randomization allocation or data that may break blind during the trial. The pragmatic nature of this adaptive platform trial increases external validity. However, we will collect information about contamination (being prescribed study therapeutics outside the trial) and co-interventions (e.g. use of inhaled steroids). We will mitigate against attrition bias by ensuring that participants are compensated for answering outcome questions regardless of adherence to the study protocol.

Randomization will occur on the day of enrollment using an interactive web-based system that will be maintained by our data management group. Participants will be allocated to a trial arm using fixed, equal randomization ratios corresponding to the number of eligible arms in the trial. Initially, patients will be randomized 1:1 to usual care and the first therapeutic to be evaluated (e.g. nirmatrelvir/ritonavir (Paxlovid™)). If a second therapeutic is added (e.g. fluvoxamine), the allocation ratio will change to 1:1:1. Patients will be stratified based on age (<65 years vs. older) ^41,42^ and will use random sized permuted blocks.

#### 5.4 **Drug description**

As this trial progresses, therapeutics for SARS-CoV-2 in out-patient settings to be evaluated will be determined by the Canadian COVID-19 Out-Patient Therapeutics Committee, which will evaluate the latest evidence on new therapeutics and makes recommendations to the Steering Committee. This approach is modelled after the UK COVID-19 Therapeutics Advisory Panel, which made recommendations to national platform clinical trials including RECOVERY, REMAP-CAP, and PRINCIPLE, among others. Following other APTs, such as I-SPY 2^43^ and STAMPEDE^44^, the criteria to decide on whether to include a new therapy will include:

- Sound scientific and biological rationale or compelling evidence for treatment of SARS-CoV-2 in out-patient settings, and related, successful independent study (phase I/II)
- Sufficient supply of the therapeutic
- Sufficient scalability of the therapeutic if found to be effective
- Adding a new arm does not jeopardize completion of ongoing research arms
- The new comparison must be relevant when it is completed

Each therapeutic to be evaluated to the trial will be added to the Master Protocol as a sub-protocol. Details on each therapeutic will include dosage, side effects and adverse reactions, duration of treatment, and existing information, along with a patient handout in lay language.

As noted in our exclusion criteria [Section 6.2], we will mitigate contamination as potential participants will be screened for current use of a trial therapeutic, and part of the recruitment process [Section 8] will include a pharmacist review to identify potential interactions with existing therapeutics. A study pharmacist(s) will be hired in each province. Prior to being consented to the study, a participant will be contacted by a research assistant for initial screening, which will include eligibility criteria and a crude review of medications, based on a list provided by the patient. Following this, the participant will review the informed consent form (ICF) with the research assistant and then consent to the study if willing to participant [Section 9]. At this point, the study pharmacist will conduct a detailed review of the participant’s medications, a process which will include obtaining a medication list from the participants usual pharmacy via verbal communication, if available, to confirm their eligibility to participate in the study. Medications will be recorded in a medication log (Appendix 9). In circumstances where the pharmacist requires more data to establish eligibility, they will examine the participant’s medical history, medication, and lab results. They will leverage relevant electronic clinical resources, such as the Connecting Ontario portal for Ontario residents, to acquire necessary information for eligibility determination. If there is still a need for more information, the study team may contact the participant's primary care provider to acquire additional details.

Following a thorough review of the participant's medications, and once their eligibility for the study is confirmed, the study pharmacist will make a recommendation to the provincial principal investigator (PI) regarding the participants eligibility. If in agreement, the provincial PI will sign off on the participants eligibility.

Once eligibility has been confirmed, participants will be randomized to a study arm and if applicable, a notification will be sent to a provincial study pharmacy to ship the study therapeutic to the patient. However, the study drug may also be securely stored within the designated space of the study team and, if necessary, be distributed to the participant by a member of the research team. This approach has been already successfully used in a variety of Canadian outpatient trials.

***Duration of treatment:*** Treatment regimens will be decided by the Canadian COVID-19 Out-Patient Therapeutics Committee based on doses and durations studied in past trials and the latest evidence. These details will be outlined in intervention specific sub-protocols.

***Compliance:*** Compliance will be captured through self-reported daily diaries up to day 14, and the 21- and 28-day follow-up surveys. ^31^ Compliance and early discontinuation may be a key issue, particularly with potentially high rates of discontinuing some therapeutics (e.g., nirmatrelvir/ritonavir early)^34,45,46^ and will be studied and reported as part of this trial.

#### 5.5 **Study duration**

CanTreatCOVID is designed as an APT, supporting ongoing research on out-patient therapeutics for SARS-CoV-2 infection. This platform allows for the study to be perpetual, with multiple treatments evaluated at any given time, and over time. Frequent adaptive analyses are performed to determine whether the interventions under evaluation are still eligible for further testing or if randomization should be stopped due to demonstrated inferiority, superiority, or equivalence. The only limit on the duration of a platform trial is the availability of ongoing funding, the availability of new interventions to evaluate, and that the disease continues to be of clinical and public health importance. Participants’ last enrollment date will be March 31^st^, 2025. The last follow up point for all participants is April 28^th^, 2025 which will ensure all participants will complete the 21 day and 28 day follow up. Participants scheduled to be followed up after April 28^th^, 2025, will not be followed up however, they will receive the total honoraria of $120 despite not completing the remaining follow ups. This will be applicable for participants who were enrolled in the study after August 19^th^, 2024, who will be unable to complete their Week 36 follow-up visit, and participants enrolled after January 28^th^, 2025, who will be unable to complete their Day 90 and Week 36 follow-up visits, prior to April 28^th^, 2025.This will be communicated by Hub staff to all patients who have follow-up visits scheduled past April 28^th^ 2025.

#### 5.6 **Study stopping rules/termination**

It is anticipated that after inclusion of the initially planned sample size, the study would continue to include additional participants and test additional domains and/or interventions until one of the following occurs:

- SARS-CoV-2 is no longer deemed to be a significant public health problem in the jurisdiction that is hosting and supporting this APT
- The effectiveness and/or cost-effectiveness of plausible interventions are known and there are no new plausible interventions to test.

#### 5.7 **Data collection and records**

The Master Linking Log (Appendix 1), ICF (Appendix 2) and demographic data (Appendix 3.1) will be maintained within each provincial hub (i.e., will not be transferred outside of provinces).

***Data collection*:** At baseline, we will collect data on sociodemographic, including date of birth, sex assigned at birth, gender identity (on a voluntary basis), education level, household income, income source, ethnicity, rurality. We will also collect health care numbers, all phone numbers, an emergency contact, an email address, and seek permission to use text messaging, as well as alternative contacts and caregivers (Appendix 3.1) for obtaining follow-up data should the need arise.

Participants in all arms will complete an online daily diary each day for **14 days**, a validated approach from similar Canadian trials.^31,34^ The daily diary will take approximately 30 minutes to complete. Participants will self-report symptoms and severity, as well as contacts with health services (e.g., hospital admissions, ED visits, outpatient visits to specialists and primary care), which will be corroborated with administrative data when the data becomes available (Appendix 3.7a). Moreover, 10% of included participants will be randomized to complete additional Flu Pro Plus questions in the daily diary, which collect detailed information on severity of symptoms (Appendix 3.7b). Of note, our primary study outcome will be reported by the patient or their alternative contact and will not rely on administrative data for publication and dissemination of timely results.

Participants will be prompted to complete online follow-up surveys using REDCap Cloud© electronically through automated emails sent through the survey software. Research staff will call participants with no internet access, as well as those who prefer to complete surveys over the phone. Each study survey will take approximately 30 minutes to complete. Participants will be counselled to take breaks by stepping away from the computer or asking the research staff to call back at a time convenience to both, as necessary and needed. Participants who have not completed their diary for at least two consecutive days before day 7 or again before day 14 will be contacted by staff by email, phone, or text. Staff will make 3 attempts over 3 days before considering it missing data.

Research staff will contact participants at 21 days and 28 days, focusing on the primary outcome, and at 90 days and 36 weeks from randomization, with a focus on post-acute sequelae of SARS-CoV-2. EQ-5D-5L will be administered at baseline, 21 days, 28 days, 90 days, and 36 weeks (Appendices 3.2, 3.3, 3.4, 3.5, 3.6). Participants’ last enrollment date will be March 31^st^, 2025. The last follow up point for all participants is April 28^th^, 2025 which will ensure all participants will complete the 21 day and 28 day follow up. Participants scheduled to be followed up after April 28^th^, 2025, will not be followed up however, they will receive the total honoraria of $120 despite not completing the remaining follow ups. This will be applicable for participants who were enrolled in the study after August 19^th^, 2024, who will be unable to complete their Week 36 follow-up visit, and participants enrolled after January 28^th^, 2025, who will be unable to complete their Day 90 and Week 36 follow-up visits, prior to April 28^th^, 2025. This will be communicated by Hub staff to all participants who have follow-up visits scheduled past April 28^th^ 2025. As a result of these missed follow-up visits (90 Days and/or 36 Weeks), data collection will be limited from these participants in regards to the secondary outcome focusing on post-acute sequelae of SARS-CoV-2.

We will apply for ongoing funding after year 1 to continue collecting quality of life data at 12 and 24 months, to support our cost-utility analysis [Section 17]. Adherence to the study therapeutic will be assessed with self-report. Participants who consent to allowing access to administrative data on health service use will be followed up to 24 months post-randomization.

***Data management:*** CanTreatCOVID will be supported at the national level through the Applied Health Research Centre (AHRC). We will access the necessary infrastructure for this trial, including data collection software through industry-standard database software (e.g., REDCap©) and secure server facilities. AHRC is a designated Research Centre of the Ontario SPOR Support Unit (OSSU) deemed suitable to provide the enabling infrastructure, scientific knowledge, and technical support required to conduct patient-oriented research, and currently supports over 50 CIHR and National Institutes of Health-supported clinical trials, registries and observational studies. Each province will have a “hub” with dedicated research staff, and recruitment will occur through “spokes” in primary care centres and other community settings, as well as other recruitment methods described [Section 8]. We will engage the expertise and staff in each participating Primary Care Practice-Based Research Network (PBRN) participating in CanTreatCOVID, as directors of each are part of the study team.

### 6. SELECTION AND WITHDRAWAL OF PARTICIPANTS

#### 6.1 **Inclusion criteria**

Inclusion: Age **50 years and older or 18-49** with 1 or more **chronic high-risk medical conditions, and/or immunosuppression**. The following is based on the PANORAMIC trial from the UK^36^, a very similar adaptive platform trial evaluating SARS-CoV-2 therapeutics in community settings: chronic respiratory disease (including COPD, cystic fibrosis and asthma requiring at least daily use of preventative and/or reliever medication); chronic heart or vascular disease; chronic kidney disease; chronic liver disease; chronic neurological disease (including dementia, stroke, epilepsy); severe and profound learning disability; Down’s syndrome; diabetes (Type 1 or Type 2); immunosuppression: primary (e.g. inherited immune disorders resulting from genetic mutations) or secondary due to disease or treatment (e.g. sickle cell, HIV, cancer, chemotherapy); solid organ, bone marrow and stem cell transplant recipients; morbid obesity (BMI >35); severe mental illness; care home resident. These align with guidelines from the Public Health Agency of Canada (PHAC)^45^, Ontario^24^, Quebec^47^, British Columbia^48^, Alberta^49^ and the Centers for Disease Control and Prevention^50^, which has maintained a list of conditions associated with greater risk of severe outcomes based on systematic reviews and observational studies. Participants must have a **positive SARS-CoV-2 test** (PCR or RAT) with proof of a positive test provided via a picture of the result, and be enrolled **within 5 days of onset of symptoms** associated with SARS-CoV-2 infection^12,25^ (1 or more of: fever or chills, cough, shortness of breath, decreased or loss of taste or smell, runny nose or nasal congestion, headache, fatigue, sore throat, muscle aches or joint pain, gastrointestinal symptoms).

#### 6.2 **Exclusion criteria**

Exclusion: Admitted to hospital or in an ED for more than 24 hours, previously randomized to CanTreatCOVID, currently participating in a clinical trial of a therapeutic agent for acute SARS-CoV-2 infection that is not/suspected not compatible with the study therapeutics, already taking a study therapeutic or contraindication to a study therapeutic, or inability for participant or caregiver to provide informed consent.

#### 6.3 **Reasons for withdrawal**

Every participant (or their legal representative on behalf of a participant) retains the right to withdraw from CanTreatCOVID at any time. For those that lack capacity, an expression of dissent that takes any form will be judged as an indication that they do not wish to be included and that participant will be withdrawn.

In addition, the investigators may discontinue a participant from CanTreatCOVID if it is considered necessary, for reasons including:

- Ineligibility (either arising during the trial or retrospectively)
- Withdrawal of consent

The reason for withdrawal will be recorded on the participants eCRF. Data that has already been collected about the participant will be kept and used. If a participant does withdraw from the study, there is no follow-up required.

### 7. POTENTIAL BENEFITS, RISKS AND SAFETY

#### 7.1 **Potential benefits**

Participants in CanTreatCOVID will be randomized to usual care or one of the treatment groups. The treatments being studied may or may not be of direct benefit. We hope this study will advance scientific knowledge to help people infected by COVID-19 in the future.

#### 7.2 **Potential risks**

Participants in CanTreatCOVID may experience side effects from participating in this study. Some side effects are known and will be listed in therapeutic specific sub-protocols, as well as in the patient handout. There may be other side effects that are not expected, and we will ensure participants are aware of how to report these and seek assistance.

Participants will be monitored closely (through online daily diaries for 14 days) to see if they have side effects. When possible, other medicine can be given by participants usual source of primary care to make side effects less serious and more tolerable. Many side effects go away shortly after the study intervention is stopped, but in some cases side effects can be serious, long-lasting, permanent, or may even cause death.

Participants will be informed that risks and side effects related to the experimental interventions and the likelihood of having the risks and side effects may be different. New therapeutics may have serious side effects not yet discovered, or long-term effects that are unknown. It is possible that other drugs (prescription and non-prescription drugs), vitamins, or herbals can interact with the study intervention. This can result in either the intervention not working as expected or result in severe side effects. As noted previously [Section 5.4], a study pharmacist will conduct a detailed review of the participant’s medications, a process which will include obtaining a medication list from the participants usual pharmacy, if available, to confirm their eligibility to participate in the study.

Reproductive risks: We anticipate that for some or all of the study therapeutics that are included, the risks on an unborn baby (fetus) are unknown. Participants will be counselled to not become pregnant or father a baby while taking the therapeutic, and for 14 days post intervention. Research staff will be trained to discuss family planning with participants to ensure that they do not become pregnant or father a baby during the study. Participants will be counseled to discuss these risks with sexual partners.

Breastfeeding: Participants will be counselled to avoid breastfeeding while taking the therapeutic and for 14 days after the last dose because the therapeutics used in this study might be present in breast milk and could be harmful to a baby.

#### 7.3 **Safety**

#### 7.3.1 ***Overview of principles regarding safety***

Research oversight – particularly for any trial that involves vulnerable populations – is essential to protect participant safety and rights and ensure public trust in how the trial is conducted to safeguard the welfare of all participants. We outline the definition, attribution, and reporting of adverse events and serious adverse events (SAEs) to achieve these goals, and to avoiding the reporting of events that are likely to be part of the course of illness.

Any therapeutic that we will assess in this adaptive platform trial would only be selected for inclusion if there was a reasonable balance of potential effectiveness to potential harms. One role for the DSMC (Section 15) will be to ensure this balance is met. We have budgeted for pharmacists in provincial hubs to be available to review potential interactions with existing medications. All participants will be called within one day of starting a therapeutic and will have access to the toll-free study hotline maintained by study staff between 7 AM to 8 PM ET. Emergency procedures will be emphasized. CanTreatCOVID will assist with obtaining important safety data that is not possible to obtain without large-scale randomization, sharing safety data in coordination with other ongoing studies.

#### 7.3.2 ***Adverse event reporting and definition***

An Adverse Event (AE) is defined as any untoward medical occurrence in a participant that may arise during or following participation in a clinical study, regardless of whether it is directly related to the study drug or intervention. This includes any worsening or deterioration of an existing clinical condition. AEs must be promptly reported and documented in accordance with the following procedures:

- Any clinical event, or worsening of a pre-existing condition, that occurs after a participant’s inclusion in the study must be reported to the Site Investigator and recorded in the case report form (CRF).
- Participants should be informed of the importance of reporting any physical changes to the research team.
- Research coordinators are responsible for accurately documenting all reported AEs in the Adverse Event electronic Case Report Form (eCRF).
- For each AE/ADR, the following information must be recorded in the eCRF: the date of onset, the date of resolution, a description of the event, intensity, outcome, actions taken, and the relationship to the study drug.

Severity of adverse events should be classified as follows:

- **Mild**: Easily tolerated by the participant, causing minimal discomfort and not interfering with everyday activities.
- **Moderate**: Sufficiently discomforting to interfere with normal everyday activities.
- **Severe**: Prevents normal everyday activities or requires significant medical intervention.

The Qualified Investigator must assess AEs based on three key factors:

- **Intensity:** The event is categorized as mild, moderate, or severe. Note that the severity of the event does not always correlate with its seriousness. For example, a severe headache may not require immediate reporting to regulatory authorities.
- **Relationship to Study Medication:** The relationship between the AE and the study drug must be assessed. Based on clinical judgment, the AE may be classified as certainly, likely, possibly, or unlikely related, or as unrelated to the study drug.
- **Seriousness:** Events are classified as serious if they involve life-threatening conditions, require inpatient hospitalization or prolongation, result in significant disability or incapacity, or lead to birth defects or congenital abnormalities.

At baseline, the research coordinator will inquire if the participant has any pre-existing medical conditions or comorbidities. These must be recorded in the *Comorbidity eCRF*, clearly indicating that they were pre-existing conditions.

During follow-up, either through surveys or phone calls, the research coordinator will inquire about new clinical experiences, diagnoses, or the worsening of existing conditions. Any changes must be recorded in the Adverse Event eCRF.

#### 7.3.3 ***Serious adverse events reporting and definition***

We define SAE as an event or reaction to the study medication that is fatal, life-threatening, requires inpatient hospitalization or prolongation, results in disability that is persistent and significant, or results in a birth defect or congenital abnormality. Any other important medical event that may not result in one of the above outcomes, may be considered a SAE when, based upon appropriate medical judgment, the event may jeopardize the participant and may require medical or surgical intervention to prevent one of the outcomes listed above.

Any SAE that might have reasonably occurred (from enrollment until week 36 follow up or until April 28th, 2025) as a consequence of a study intervention or study participation will be reported by trial staff to the PI and entered in an adverse event log (Appendix 8) within 24 hours of trial staff becoming aware of the event.

For participants that will be monitored until April 28th, 2025, the participant will be followed up for a total of 28 days. The antioxidant investigational product total course of treatment is 10 days, and the Investigator Brochure informs us that potential adverse effects occur primarily during the 10 days treatment course. Thus, a total of 28 days of participant follow up will be sufficient to monitor participant safety as it relates to the investigational product.

For participants that were enrolled during the Paxlovid treatment arm, there are no concerns regarding missing follow up visits with respect to Adverse Drug Reaction monitoring. The investigational product course of treatment is 5 days, where the majority of adverse drug reactions occur during or shortly after the treatment course is complete

The study sponsor will be notified within 72 hours. The minimum information that will be reported will comprise:

- Unique participant ID
- Date(s) of the event
- Nature of the event, including the outcome and rationale for attribution to a trial intervention
- Whether treatment was required for the event, and, if so, what treatment was administered

SAEs that are serious, unexpected and likely have a drug-related cause are subject to expedited reporting to Health Canada. An adverse drug reaction (ADR) report will be filed in all cases:

- where the ADR is neither fatal nor life-threatening, within 15 days after becoming aware of the information
- where it is fatal or life-threatening, immediately where possible and, in any event, within 7 days after becoming aware of the information

Within 8 days after having informed Health Canada of the ADR, we will submit as complete as possible, a report which includes an assessment of the importance and implication of any findings.

SAEs will be monitored from the time of randomization to week 36 or until April 28^th^, 2025. SAEs may be collected through the daily diary, communicated on study surveys or to research staff directly, or communicated through the toll-free study hotline (1-888-888-3308) maintained by study staff between 7 AM to 8 PM ET. Patients will be follow-up with regarding the SAE until external medical care is sought or until resolution.

***Attribution of SAEs to study interventions***

It may be difficult to distinguish, in real-time, between events that occur as a consequence of illness (i.e., infection with SARS-CoV-2), treatments not specified by the trial, and interventions specified by the trial. The standard that will be applied to determine whether SAEs are attributable to study interventions in this trial is that it is unrelated, unlikely, possible, probable, or definite that there is a direct link between a trial intervention and the SAE, or the SAE is not considered to be a normal feature of the evolution SARS-CoV-2 infection.

The primary outcome of CanTreatCOVID is all-cause hospitalization and/or death at 28 days, and the objective of this study is to identify differences in this primary outcome that can be attributed to treatment allocation, given that it is not known whether some treatments are more effective than others. As this trial evaluates treatments that are not necessarily part of the usual standard of care, the threshold for considering attribution of SAE or death to the novel treatment will be lower than if the treatment was already in widespread use and its safety profile already established.

### 8. RECRUITMENT

We will recruit patients using multiple methods.

***1) Public communications and community organizations:*** All recruitment materials will be in English and French, with additional translations in up to the top five non-English/French languages in each province. We will use a website (www.CanTreatCOVID.org) with clear information for the public to engage directly with the study, building off lessons learned from prior Canadian trials as well as PANORAMIC in the UK. This will also support transparency and knowledge mobilization. We will engage a professional public communications firm to create recruitment materials (see Appendix 4.1), for public advertisement (transit advertisements, local print/online media, traditional media, email/mail/SMS/fax marketing) and social media (Google ads, Facebook, Twitter, Instagram, TikTok, and other online channels). We will work with local community organizations in key participating cities to disseminate broadly to diverse communities. We have established a toll-free study hotline (1-888-888-3308), active from 8AM to 6PM ET Monday- Friday to support ongoing access, ensuring our team of research assistants are available to intake participants and address questions.

***2) Prospective engagement through primary care practice-based research networks:*** Study team members include national and international experts in using primary care electronic medical data (EMR) data for trial recruitment. We will engage practices and physicians, and with their permission, will use existing primary care EMR data from these networks to identify patients who meet our eligibility criteria. This would then allow prospective communication to facilitate enrollment should they become infected as described below. This has been effective in recruitment for other trials we have led. In clinics where physicians act as their own Health Information Custodian (HIC), we will seek permission (Appendix 4.4) to request access their patient EMR data for use in the study. In clinics where a designated (HIC) is assigned for an entire clinic, we will request permission from the Central HIC for the clinic as a whole to participate (Appendix 4.4.1). After receiving permission from the Central HIC, a two week-opt out period will commence where each individual physician is emailed with the opportunity to leave the study.

Family physicians and nurse practitioners will be invited to take part in the study. If family physicians and nurse practitioners agree, we will **search primary care EMR data for patients** that are 50+ years old or 18-49 years old with 1+ chronic medical condition and/or who are immunosuppressed (see inclusion criteria in Section 6.1). In cases where we are not granted direct access to the EMR data, we acknowledge the administrative time commitment this process entails. As such, we are prepared to reimburse any staff time involved in this data access at the rate of $40/hr. This is in recognition of the valuable role the administrative staff play in this research and to minimize any potential disruption to their routine tasks.

Anticipating that a certain proportion of the population will be infected with SARS-CoV-2 during times of increased transmission, with family physician and nurse practitioner agreement, we will proactively reach out to eligible participants (via mail or email) on behalf of their care team, notifying them of the study (Appendix 4.5) and how to get in touch if symptomatic. Where access to RATs had been limited, we may also provide free RATs. In addition, for providers who prefer to send mass communications instead of individual letters, we will send bulk letters (through email or mail) to potentially eligible patients (Appendix 4.5.1).

In addition, these networks allow us to disseminate information on the trial to hundreds of primary care providers, to ensure they consider referring patients with SARS-CoV-2 to CanTreatCOVID.

**3)** ***Outreach through various out-patient setting:*** Our team includes primary care and specialist clinicians based in numerous out-patient settings, including but not limited to community primary care clinics and health centers, urgent care clinics, specialty clinics, as well as infectious disease clinics, emergency departments, and pharmacies. We will disseminate recruitment materials (e.g., posters) broadly, and also use internal communication channels to regularly remind providers about referring patients, in particular to providers for high-risk out-patient populations such as oncology and transplant patients. Additionally, a $40 referral fee will be provided to healthcare providers and pharmacists for referring patients who meet the key study inclusion criteria and have passed the initial screening with research assistants. Importantly, this fee will be provided regardless of whether or not the potential patient decides to consent to participate in the study. This approach, as corroborated by its successful implementation in similar studies such as PANORAMIC^51^, highlights the critical role of healthcare professionals in enhancing the success of research studies.

***4) COVID-19 Assessment Centres:*** Although changes in criteria for PCR testing has greatly limited the number of tests done, this may change in the future. In each jurisdiction, working with knowledge users and policymakers, we will pursue a process to automatically notify those who have a positive result about the study, and how to take part.

***5) Leveraging institutional and provincial databases for participant engagement:*** Our interdisciplinary team is affiliated with various institutions, all of which maintain databases of participants from past studies who have consented to future contact for research purposes. This valuable resource allows us to directly connect with potential participants who have previously demonstrated a willingness to contribute to scientific research. In addition to our internal databases, we will also collaborate with provincial organizations such as the Ministry of Health. Our goal is to gain access to their database of patients who have received the COVID-19 vaccination and have given their consent to be contacted for research purposes. Of course, this strategy is contingent on receiving the necessary regulatory approvals from these organizations. We will work closely with these entities to ensure all data sharing complies with the relevant privacy and ethical guidelines. Once we have gained the necessary approvals and access to the databases, our team will reach out to potentially eligible participants via mail, SMS, or email, depending on the contact preferences they have indicated. We will provide them with information about the study (as detailed in Appendix 4.5.2) and how to reach out if they become symptomatic.

6) Engaging with Contract Research Organizations (CROs) for participant recruitment: Recognizing that recruitment can be a challenge, we will engage CROs to identify eligible participants for the CanTreatCOVID study. The CROs will employ a diverse range of recruitment strategies to identify potentially eligible participants during the initial screening phase and will facilitate contact between the participants and the study team for enrollment. In addition to supplementing existing social media posts and advertising, the CROs will provide insights and feedback to enhance recruitment strategies further. Leveraging both their internal and external networks, the CROs will actively promote study materials. Their involvement in recruitment and initial screening will be essential to the process, as detailed in Appendix 14.

### 9. CONSENT AND SCREENING

The consenting process for the trial will be completed by a research assistant, who is unknown to the potential participant, over the phone using our information sheet and ICF (Appendix 2). Through one of the recruitment methods [Section 8], potential study participants will make initial contact with a research assistant either electronically via email or through a provided phone number to begin the enrolment process. Recruitment communications will advise potential participants to contact the study as soon as possible from symptom onset to ensure that screening, consenting, and shipment of study medications can take place within 5 days of symptom onset. During the initial contact, the research assistant will describe the study and conduct eligibility screening, including a crude review of medications reported by the potential participant to ensure none are contraindicated to the study medications being investigated. If the participant agrees to participate in the trial, we offer three methods to provide their consent:

1. E-Consent Module: Participants can use the E-Consent module on the Research Electronic Data Capture (REDCap) system, hosted on the secure servers at the Applied Health Research Centre (AHRC) at St. Michael’s Hospital, to provide consent. Once participants opt for this method, they will receive an email containing a link to the ICF. Upon clicking the link, participants will be directed to a secure webpage where they can review the ICF at their own pace. After reviewing the ICF, if the participant agrees to participate in the study, they can provide their consent by signing the online form. All information provided through REDCap is encrypted and securely stored, ensuring the highest degree of confidentiality and data protection.

2. Email Consent: Participants can receive the ICF and return a signed copy of the ICF via email.

3. Verbal Consent: If options 1 and 2 are not feasible, verbal consent will be sought via telephone, in the presence of a witness. The witness will also provide an attestation (Appendix 2.1) that the potential participant has provided their informed consent.

To ensure comprehensive eligibility assessment, the provincial study pharmacist will review the provided list of medications, which will include obtaining a medication list from the patient’s usual pharmacy through verbal communication. For cases where additional information is needed to assess eligibility, the study pharmacist will review the patient's medication, medical history, and lab data. They will access appropriate electronic clinical resources, such as the Connecting Ontario portal for participants reside in Ontario, to gather the necessary information and determine eligibility. Only if additional information is still needed, the study team may reach out to the patient's primary care provider to obtain further information. Once the review of medications is complete and eligibility for the study is confirmed, the pharmacist will recommend to the PI in each province to approve or not approve the participant for the study. If the provincial PI agrees that the participant is eligible for the study, the participant will be randomized to a study arm, shipped the study medication if applicable, and data collection will begin.

The following messages will be emphasized during the consenting process: 1) Participants can take up to two days to make their decision and can discuss it with friends and family (this is necessary to ensure that study medications can be taken within 5 days of symptom onset, the timeframe in which they are most effective), 2) Taking part in this study is voluntary, and participants have the option to not participate at all or leave the study at any time without penalty, and 3) Participants’ decision will not affect the usual medical care that they receive outside the study. Research staff will be trained to confirm ongoing consent at the beginning of each study encounter.

As noted in the consent form, we will also confirm consent to notify the participant’s primary care provider (if one exists) about participation in the trial. therapeutic specific sub-protocols will include a template letter to primary care providers about the specific intervention that the participant has been randomized to. While we will carefully review drug interactions with our study pharmacists, this notification will act as a second safety check, and keep the primary care provider updated on the status of the participant. The schedule of events for participants is described in Table 1.

***Table 1: Participant schedule of events***

| **Event** | **Description** |
| --- | --- |
| Initial Contact | A potential participant is connected with a research assistant through one of the recruitment methods described [Section 8]. |
| Screening | Participant is contacted by a research assistant for initial screening, including eligibility criteria and a crude review of medications. |
| Consent | Participant reviews the ICF over the telephone with the research assistant and is sent the ICF electronically via email, if possible. If willing to participate, the participant will verbally consent to be a part of the study and if possible, will electrically return a signed copy of the ICF via email. |
| Pharmacist’s assessment of eligibility | A study pharmacist (or site investigator) will perform a comprehensive review of the participant's medication list, which may involve obtaining additional data from the participant's regular pharmacy via phone. If additional information (medical history, lab data, etc.) is needed to assess eligibility, pharmacist may also use relevant digital health resources, like the Connecting Ontario portal for Ontario-based participants, to gather the necessary information. In cases where further details are required, the study team might reach out to the participant's primary care provider. The study pharmacist will then provide their recommendation to the PI regarding the participants eligibility for the trial. |
| Participant eligibility confirmed | If both the provincial study pharmacist and PI agree regarding the medication review, the participant is enrolled in the trial. |
| Randomization | Participant is randomized to a trial arm. |
| Baseline data collection | Participant completes baseline data collection electronically or over the telephone with a research assistant to provide information on sociodemographic characteristics, vaccination status, time to recovery, health service use, and quality of life. |
| Medication order placed | If participant randomized to a treatment arm, either the distribution pharmacy receives the order and ships the study drugs or they may be stored and shipped by the study team from a secure location. |
| Day 1 check-in | Research assistant connects with the participant to confirm that the medication was received and started within the 5 day timeline and that there were no serious adverse events upon taking the first dose. |
| Daily diary (14 days from onset of study medication use) | Participant completes an online daily diary for 14 days, assessing time to recovery, symptoms, symptom severity, continuation of study medication, other medication use, and healthcare services use. |
| Follow up at 21 days | Participant completes the 21 day follow up electronically or over the telephone with a research assistant to provide information on time to recover, health service use, early discontinuation, and quality of life. |
| Follow up at 28 days | Participant completes the 28 day follow up electronically or over the telephone with a research assistant to provide information on time to recover, health service use, early discontinuation, long COVID, and quality of life. |
| Follow up at 90 days | Participant completes the 90 day follow up electronically or over the telephone with a research assistant to provide information on time to recover, health service use, long COVID, and quality of life until April 28^th^, 2025. Participants with this visit scheduled after April 28^th^, 2025, will not complete this follow up. |
| Follow up at 36 weeks | Participant completes the 36 week follow up electronically or over the telephone with a research assistant to provide information on time to recover, health service use, long COVID, and quality of life until April 28^th^, 2025. Participants with this visit scheduled after April 28^th^, 2025, will not complete this follow up. |
| Follow up at 12 months | Review of administrative data to determine health service use, if participant consented to allowing access to data. Pending additional funding after Year 1, quality of life data will be collected at 12 months and converted using standard approaches to QALYs. Cost-utility analysis will be calculated as the incremental cost per QALY gained until April 28^th^, 2025. |
| Follow up at 24 months | Review of administrative data to determine health service use, if participant consented to allowing access to data. Pending additional funding after Year 1, quality of life data will be collected at 24 months and converted using standard approaches to QALYs. Cost-utility analysis will be calculated as the incremental cost per QALY gained until April 28^th^, 2025. |

### 10. SPECIMEN COLLECTION, STORAGE AND ANALYSIS

Not applicable, as CanTreatCOVID will not include the collection of specimens.

### 11. INCENTIVES AND COMPENSATION

Participants will be offered $30 CDN for each study encounter (e.g., at 21 days, 28 days, 90 days, 36 weeks, etc.). For those who have follow ups scheduled after April 28^th^, 2025, (for example 90 days and 36 weeks) will still receive compensation for a total of $120. This will be applicable for participants who were enrolled in the study after August 19^th^, 2024, who will be unable to complete their Week 36 follow-up visit, and participants enrolled after January 28^th^, 2025, who will be unable to complete their Day 90 and Week 36 follow-up visits, prior to April 28^th^, 2025.

### 12. INCIDENTAL FINDINGS

Given the nature of CanTreatCOVID, which is focused on short-term therapeutics for SARS-CoV-2 infection, with follow-up using a combination of self-reported daily diaries, and electronically- or research staff- administered surveys at baseline, 21 days, 28 days, 90 days, and 36 weeks, we do not anticipate many, if any, incidental findings until April 28^th^, 2025. This study does not include any specimen collection, imaging, or diagnostic testing, so incidental findings will not be discovered via these avenues.

In the event an incidental finding is discovered through follow up, the participant will be contacted by research staff via telephone and advised to get in touch with their primary care provider. During the consenting process, participants will be asked permission for research staff to contact them and/or their primary care provider if such events occur, and related contact information will be collected during baseline data collection.

In the case where a participant does not have a primary care provider, research staff in each of the participating provinces will be equipped with information on how participants can use provincial resources to find a primary care provider. Our study team includes primary care physicians in each of the participating provinces, who can serve as a final resource to support participants with incidental findings. In the consenting process, participants will also be asked permission for provincial study investigators to follow up with them regarding any incidental findings in the absence of a primary care provider.

We will also ensure that each participating provincial hub has a list of local centres who specialize in long COVID such that patients who identify as having this disease, or who are identified, can seek appropriate care.

### 13. PROTOCOL DEVIATIONS

Analyses will be carried out in accordance with the Master Protocol and Statistical Analysis Plan. No deviations from this protocol will be permitted without the prior written approval of the Sponsor, except when the modification is needed to eliminate an immediate hazard or hazards to participants. Any deviations that may affect a participant’s treatment or informed consent, especially those increasing potential risks, must receive prior approval from the REB unless performed to remove an immediate safety risk to the participants. In this case it will be reported to the REB and the Sponsor immediately thereafter. Any departures from the protocol will be documented (Appendix 11).

**Minor protocol deviations** (i.e. out-of-window visits for follow-up) or **Major protocol deviations** (i.e. improper consent procedure, missing COVID + test result) should be documented appropriately on the Protocol Deviation Log **within 7 days** and if required reported to the Sponsor-PI team and REB accordingly.

All Protocol Deviations should be reviewed by the Qualified Investigator on a monthly basis.

Any Protocol Deviation identified later than 7 days of its occurrence in addition to documenting in the Protocol Deviation Log should also be documented in a Note to File detailing the reason for delayed reporting. This NTF should be signed off by the Qualified Investigator and shared with the site team about corrective and preventative actions to avoid future deviations.

### 14. CONFLICT OF INTEREST

The Principal Investigator declares no potential conflict of interest. All study team members for CanTreatCOVID will be required to complete a detailed disclosure of conflicts of interest, and these will be posted on our open website.

### 15. DATA MONITORING

***Data Safety and Monitoring Committee:*** The DSMC will include a chair and 4 additional members that are external to the study team (Appendix 5: DSMC Charter; Appendix 7.2: Committee members). All DSMC members for CanTreatCOVID will be required to complete a detailed disclosure of conflicts of interest, and these will be posted on our open website. The DSMC will serve a crucial role, receiving results reported as Group A (arm 1), Group B (arm 2), etc. Should there be a significant difference in outcomes or adverse events, data will be unblinded to the DSMC chair and then shared with the Steering Committee to reach a unanimous decision with respect to next steps (i.e., continue or stop the trial). Stopping rules will be pre-determined and included in the Statistical Analysis Plan related to efficacy, futility, and safety.

### 16. KNOWLEDGE TRANSLATION AND PUBLICATION

In keeping with the principles of integrated knowledge translation, as outlined by the CIHR, those most likely to benefit from or use the knowledge produced have been involved from inception, including in the design (e.g., confirming primary and secondary outcomes, recruitment, therapeutics to evaluate), interpretation and dissemination of findings.

We will work closely with our current knowledge users, who are from the National Collaborating Centre for Infectious Diseases, Réseau-1 in Quebec, Ontario’s Office of the Chief Medical Officer of Health, Alberta’s Associate Chief Medical Officer of Health, primary care leaders, and those developing provincial COVID therapeutic guidelines. We will continue to engage knowledge users from PHAC and via our networks including through the CIHR Applied Public Health Chairs program and the SPOR Primary Health Care networks.

***Patient and citizen engagement:*** We have already engaged a number of patient partners and we have budgeted for a total of 25 patient partners from across Canada. Our patient engagement will go beyond tokenism to engage the patient perspective in a meaningful and sensitive manner, building on our experience in engaging patients directly in research study teams. Specifically, our patient and citizen advisors will provide input on the recruitment materials, act as local and national ambassadors (e.g., video clips on website explaining the study), and advise on the outcomes and test data collection tools and consent processes.

***Community engagement:*** Our team members have extensive experience working with community partners on research. We know that organizations serving communities made vulnerable by social and economic policies want to ensure treatments are accessible and based on the best evidence generated from diverse participants. We have allocated funds for a central community engagement specialist and for each provincial hub to have a community engagement lead, who will assist with building a local network of contacts, soliciting feedback to tailor recruitment materials, and to support recruitment and local knowledge translation.

Our team takes seriously the need to ensure that any research pertaining to Indigenous communities is initiated based on community priorities, maintains Indigenous leadership, and that data governance and sovereignty is upheld. We are committed to building ongoing relationships with Indigenous governance organizations, communities, and Indigenous researchers and leaders that starts by listening to communities about their priorities regarding COVID-19 out-patient therapeutics. If this is a priority, we would work towards forming relationships that ensure Indigenous communities maintain control of the process of recruitment, community engagement, access to therapeutics and data (following OCAP™ principles^52^). We have allocated funds for an Indigenous community engagement specialist.

***Clinical care networks:*** We have strong links to frontline primary care clinicians, and study team members have national roles within the infectious disease community. We will disseminate new knowledge from our project through virtual webinars with provincial primary care networks and COVID evidence tables, and publications in scholarly journals, including: 1) trial protocol; 2) primary findings from our trial; 3) economic evaluation; 4) long-term outcomes and health service resource use at 12 months and at 2 years. The results of this study will provide high quality evidence on current and emerging therapeutics for SARS-CoV-2 and directly influence standards of care for SARS-CoV-2 infection in community settings in Canada and around the world.

***National and international linkages:*** CanTreatCOVID follows a similar approach to the PRINCIPLE and PANORAMIC APTs in the UK, and members of the study team who led these studies are contributing their extensive experience. Further, our project builds on Canadian experience with APTs during the COVID-19 pandemic, including TOGETHER, REMAP-CAP and CATCO.

As virulence of future variants is unknown, the last two years of the COVID-19 pandemic show that we need to be prepared and have APTs in place. Having APT infrastructure in place in the UK to study influenza^53^ helped to quickly initiate PRINCIPLE to focus on SARS-CoV-2. CanTreatCOVID will impact on patient care, supporting Canadians to access therapeutics with a strong evidence base and to more quickly link patients to novel agents. Our findings will impact providers and clinical guidelines, and influence funding and approvals by PHAC and Health Canada. Beyond SARS-CoV-2, CanTreatCOVID will also establish APT infrastructure to study therapeutics for influenza and other upper respiratory pathogens, and for other diseases in the future.

### 17. STATISTICAL PLAN

#### 17.1  **Sample size justification**

Our main scenario, focused on the primary endpoint of hospitalization or death at 28 days. We have determined our maximum sample size per arm based on Bayesian logistic regression to estimate the adjusted odds ratio for hospitalization or death at 28 days for a pairwise comparison of treatment arm vs. comparator (initially usual care). Assuming a conservative 5% comparator event rate of hospitalization or death^53,54^, we deemed that a reduction from 5% to 3.35% would be clinically meaningful, based on the most recent published evidence.^16,31,35^ This would correspond to a number-needed-to-treat of 60. Enrolling up to 2,256 patients per arm will provide 80% power to detect 33% RR for each of the pairwise comparison at 5% CER and a two-sided type I error rate of 5% (Table 2).

We expect fewer participants to be required in a fixed two-arm setting, especially when there is either a larger treatment benefit or futility of the intervention, which our APT design would permit. During interim analyses, decision to stop or continue enrollment to an intervention arm will be based on pre-specified stopping criteria for superiority (e.g., >99% posterior probability of RR <1.0) or futility (e.g., <10% posterior probability of RR <1.0).

***Table 2: Power and sample size requirements using risk of hospitalization or death at 28 days***

| 90% power | | | 80% power | | |
| --- | --- | --- | --- | --- | --- |
| Control | Treatment | N per arm | Control | Treatment | N per arm |
| 5.0% | 3.3% | **2981** | 5.0% | 3.3% | 2413 |

We estimate that our current funds will support recruiting 10,000 patients, allowing enrollment of up to a max of **2, 500 patients in each of four intervention arms**.

#### 17.2  **Expected rate of loss to follow-up**

Due to the short duration (28 days) to the primary endpoint, and our past experience with out-patient trials (including PRINCIPLE and PANORAMIC) involving predominantly patients engaged through primary care,^57–60^ we expect minimal loss to follow-up of 5%. As described, staff will call participants with no internet access or those who have not completed their diary for at least two consecutive days before days 7 and again before day 14. We will also ask participants to provide multiple alternative contacts if they cannot be reached. In addition, health administrative data will be used to corroborate self-reported health service use.

#### 17.3  **Health services data and economic evaluation**

A prospective economic evaluation will be conducted alongside the trial following Canadian Agency for Drugs and Technologies in Health (CADTH) guidelines^61^ and ISPOR guidelines for cost-effectiveness analysis based on clinical trials.^62^ We will assess the treatment costs, the cost-effectiveness with the ICER and the cost-utility of each therapeutic tested, using the healthcare payer perspective.^61^ In addition, we will adopt the patient or informal caregivers (e.g., out-of-pocket payments, cost and time for travel, private insurance premiums) and societal perspective (e.g., lost productivity, short-term and long-term absences from work arising from patients’ inability to work, and changes in these losses and absences associated with a therapy). Final effectiveness outcomes for the economic evaluation will be determined after the consultation with patient and community partners. Costs will include treatment costs (medication costs, administration and monitoring, and other resources needed to embed treatment into routine clinical practice), other healthcare service resource use costs associated with the managing the disease (e.g. additional primary care and specialists visits, hospital admissions), and costs of managing adverse events caused by treatment., ascertained by linking participant health card number (deterministic linkage) or first and last name and date of birth (probabilistic linkage) to health administrative databases, using existing total health service cost algorithms (primary care encounters, hospital admissions, ED visits, medication costs).^61^ Unit costs will use national standards from CIHI or provincial sources to generate a total cost per participant during the full trial time horizon.^61^ Costs data will come from the trial directly, and when necessary, data will be supplemented from published sources. EQ-5D-5L data collected at baseline, 28 days, 90 days, and 36 weeks (and at 12 and 24 months, pending additional funding after Year 1) will be converted using standard approaches to QALYs, and cost-utility analysis will be calculated as the incremental cost per QALY gained^61^, with secondary analysis for the incremental cost per sick day avoided at 28 days. Costs and health outcomes will be discounted using a 1.5% rate. All costs will be reported in 2024 Canadian dollars. Multiple imputation will be used for missing data, and sensitivity analysis will be used to examine the impact of uncertainty, including varying our discount rate from 1% to 5%.^61^ We will compare results against Canadian standard cost-effectiveness thresholds for the value of an additional QALY (generally estimated at $50,000^61^). Other effectiveness outcomes will shape the Value-Based Analysis (VBA). The VBA will be conducted for medication following core principles of value-based healthcare (VBHC) where Value = Health outcomes that matter to patients/Costs of delivering those outcomes (Costs throughout the patient journey).^63^ We will use net benefit regression to examine whether ICERs change for different sub-populations (e.g., male/female, vaccination status, comorbidities, SES).

#### 17.4  **Analytic plan**

The statistical analysis plan including interim analysis will be finalized by the Methods and Statistical Analysis Committee of CanTreatCOVID (Section 19). The interim analysis plan will be determined using a series of clinical trial simulations to ensure that type I error rate can be controlled at the marginal level, while attempting to maximize the statistical power. Before any unblinded analyses are performed by the Independent Statistical Center, the simulations will be performed by the Methods and Statistics Committee who will be blinded to the knowledge of group assignment.

The primary analysis will be a Bayesian logistic regression model of the primary outcome (hospitalization or death at 28 days), stratified by age following intention-to-treat. A corresponding Bayesian posterior distribution will be derived for the estimated log odds ratio. The primary analysis for intervention *j* will test the following hypothesis: 𝐻0: 𝜃𝑗 ≥ 0; 𝐻1: 𝜃𝑗 < 0. Let 𝜃𝑗 denote the log odds ratio comparing the odds of hospitalization or death for persons in treatment group *j* versus persons in the active comparator arm.

#### 17.5  **Planned subgroup analyses**

Previous research on COVID-19 has identified biological sex as a key variable for outcomes^64,65^, and there is emerging evidence on sex differences in the prevalence of long COVID.^66,67^ Little research exists on gender and how it relates to treatment response. Building on our experience in collecting sex and gender data^68^ and the use of this sex- and gender-based data to identify inequities^69^, we will collect data on both sex and gender identity, allowing us to present disaggregated findings, and explore any differences in outcomes by sex and gender identity. In addition, we will examine intersectionality of sex and gender with other key social determinants of health, including income, race and ethnicity, and rurality. Other subgroup analyses will focus on individuals with specific medical conditions. The number of vaccine doses received, and place of recruitment. We will also report on the number of participants who required a Substitute Decision Maker (SDM), and in addition, will perform a sensitivity analysis that excludes responses provided by a SDM and compares this to the full dataset.

### 18. ACCESS TO SOURCE DATA/DOCUMENTS

Investigator/institution will permit trial-related monitoring, audits, REB review and regulatory inspection, direct access to source data/documents

### 19. STUDY TEAM AND GOVERNANCE

We have a strong and diverse study team, committed to transparency and accountability in research and the use of public funding, using consensus decision-making to guide this work in a dynamic context.

***Steering Committee:*** The Steering Committee will provide overall direction to research staff, receive reports from each committee and hub, oversee study operations, act as a central point of contact for knowledge users and collaborators, and implement knowledge translation activities.

***Methods and Statistical Analysis Committee****:* This Committee will liaise with the study statistician, and support changes and updates to the Statistical Analysis Plan.

***Recruitment and Communications Committee:*** This Committee will support the use of primary care EMR data for outreach, as well as support local practice engagement, linkage to emergency departments for recruitment, and link to infectious disease clinics and colleagues to support recruitment. This Committee will be supported by 25 patient/citizen representatives from diverse (age, gender identity, racial identity) populations in different geographic and socioeconomic contexts, as well as representative from community organizations. We have budgeted funds to support their time to provide feedback on recruitment, community engagement, and communications materials and strategies, consent forms, data collection tools and help interpret findings. Patient partners are recruited through various channels, including the CTO College of Lived Experience and their broader networks, REACHBC, and the networks of study team members.

The Recruitment & Communications Committee, including patient partners, will meet monthly, to be revised as the needs of the study evolve. Meeting minutes will be taken during each meeting and will clearly outline action items and decisions made. Minutes will be made available upon request. Additionally, a living document tracking patient partner suggestions and decisions and how and which of these were incorporated into the trial will be maintained over the course of the study.

***Canadian COVID-19 Out-Patient Therapeutics Committee***: Study team members were invited to the Committee based on expertise in infectious diseases, primary care, pharmacology, and epidemiology. We will also invite representatives from COVID therapeutic guideline developers in provinces/territories outside of our participating provinces, the Association of Medical Microbiology and Infectious Disease Canada, the College of Family Physicians of Canada, and PHAC. Recruitment, deliberations, and decisions will be transparent.

An open call was circulated through social media and investigator networks to join the Committee and candidates were asked to submit a CV and letter of interest. Applications will be reviewed by existing members of the Committee for relevance of experience.

The Committee will initially meet monthly, to be revised as the needs of the study evolve over time. Decisions regarding including, exclusion, stopping, adding, etc. therapeutics to the trial will be based on interim findings, any newly published data from Canadian or international trials, and integration of results from international trials.

Meeting minutes will be taken during each meeting and will clearly outline action items and decisions made. Minutes will be made available upon request.

49.	Alberta Health Services. COVID-19 Scientific Advisory Group Rapid Evidence Report. 2021.

50.	Centers for Disease Control and Prevention. Science brief: Evidence used to update the list of underlying medical conditions associated with higher risk for severe COVID-19 [Internet]. 2022 [cited 2022 Mar 14];Available from: https://www.cdc.gov/coronavirus/2019-ncov/science/science-briefs/underlying-evidence-table.html

51.	Butler CC, Hobbs FDR, Gbinigie OA, et al. Molnupiravir plus usual care versus usual care alone as early treatment for adults with COVID-19 at increased risk of adverse outcomes (PANORAMIC): an open-label, platform-adaptive randomised controlled trial. Lancet 2023;401(10373):281–93.

52.	The First Nations Information Governance Centre. The First Nations principles of OCAP. 2019.

61.	Canadian Agency for Drugs and Technologies in Health. Guidelines for the economic evaluation of health technologies, 4th edition. Ottawa, Canada: 2017.

62.	Ramsey SD, Willke RJ, Glick H, et al. Cost-Effectiveness Analysis Alongside Clinical Trials II—An ISPOR Good Research Practices Task Force Report. Value in Health 2015;18(2):161–72.

63.	Canadian Foundation for Healthcare Improvement. Value-based healthcare toolkit. Ottawa, Canada: 2020.

64.	Haitao T, Vermunt J V., Abeykoon J, et al. COVID-19 and Sex Differences. Mayo Clin Proc 2020;95(10):2189–203.

65.	Penna C, Mercurio V, Tocchetti CG, Pagliaro P. Sex‐related differences in COVID‐19 lethality. Br J Pharmacol 2020;bph.15207.

66.	Scully EP, Haverfield J, Ursin RL, Tannenbaum C, Klein SL. Considering how biological sex impacts immune responses and COVID-19 outcomes. Nat Rev Immunol 2020;20(7):442–7.

67.	Wolfe J, Safdar B, Madsen TE, et al. Sex- or Gender-specific Differences in the Clinical Presentation, Outcome, and Treatment of SARS-CoV-2. Clin Ther 2021;43(3):557-571.e1.

68.	Pinto AD, Aratangy T, Abramovich A, et al. Routine collection of sexual orientation and gender identity data: a mixed-methods study. Can Med Assoc J 2019;191(3):E63–8.

69.	Kiran T, Davie S, Singh D, et al. Cancer screening rates among transgender adults. 2019;65:30–7.

### 21. APPENDICES

Appendix 1: Master Linking Log and participant contact information

Appendix 2: Informed Consent Form

- 2.1 – ICF witness attestation form

Appendix 3: Questionnaires and data collection

- 3.1 – Demographic data
- 3.2 – Baseline data
- 3.3 – Follow-up visit at 21 days
- 3.4 – Follow-up visit at 28 days
- 3.5 – Follow-up visit at 90 days
- 3.6 – Follow-up visit at 36 weeks
- 3.7a – Participant daily diary (14 d)
- 3.7b – Participant daily diary (14 d) Flu Pro Plus
- 3.8 – Follow-up at day 1
- 3.9 – End of treatment end of study

Appendix 4: Recruitment materials

- 4.1 – Initial contact script & screening form
- 4.2 – Pharmacist script & pharmacy forms
- 4.3 – Recruitment poster
- 4.4 – Letter to PBRN-associated providers
- 4.4.1 – Letter to PBRN-associated HIC
- 4.5 – Letter to PBRN-associated patients from providers
- 4.5.1 – Outreach Letter, Email and SMS
- 4.6 – Letter to providers after patient enrollment
- 4.7- Online self-screening form

Appendix 5: DSMC charter

Appendix 6: Intervention Specific Appendices

- 6.1 – Intervention Specific Appendix: Usual Care

Appendix 7.1: Study co-investigators

Appendix 7.2: Committee members

Appendix 8: Adverse event log

Appendix 9: Medication log

Appendix 10: Amendment tracker for CanTreatCOVID

Appendix 11: Protocol Deviation Log

Appendix 12: Toll-free phone line contacts

Appendix 13: Deputized agent agreement

Appendix 14: Strategies and role of Contract Research Organizations in participant recruitment for CanTreatCOVID
