## Supplementary material for "Efficacy of antioxidant therapy in mild to moderate SARS-CoV-2 infection: A pilot experimental arm of the CanTreatCOVID adaptive platform trial": Antioxidant Therapy Sub-Protocol

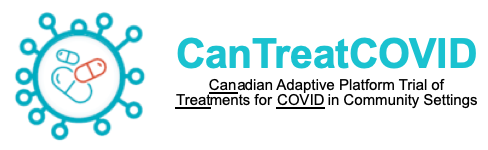

**Can**adian Adaptive Platform Trial of **Treat**ments for **COVID** in Community Settings

**Intervention Specific Sub-Protocol:** Antioxidant Therapy x 10 days

**Principal Investigators:** Benita Hosseini MSc PhD

David Jenkins MD PhD DSc

Andrew D. Pinto MD CCFP FRCPC

**Contact Details:** Upstream Lab, MAP/Centre for Urban Health Solutions, Li Ka Shing Knowledge Institute, Unity Health Toronto, 30 Bond Street, Toronto, Ontario, Canada M5B1W8,, 416-864-6060 x76148

**Study Sponsor:** Unity Health Toronto

**Contact Details:**

**Study Funder:** Canadian Institutes for Health Research (CIHR)

**Contact Details:**

**Date:** October 25, 2024

**Protocol Version:** 3.0

**Clinical Trials.gov identifier:** NCT05614349

**SITE INVESTIGATOR SIGNATURE PAGE**

Title: Canadian Adaptive Platform Trial of Treatments for COVID in Community Settings (CanTreatCOVID)

This is a sub-protocol of the Canadian Adaptive Platform Trial of Treatments for COVID in Community Settings (CanTreatCOVID). Participants meeting the platform and this sub-protocol inclusion and exclusion criteria will be randomized to receive a combination antioxidant therapy (comprising of selenium, zinc, lycopene, and vitamin C) once daily for 10 days.

I agree to the terms and conditions relating to this study as defined in this protocol. I will conduct this study as outlined and will make a reasonable effort to complete the study within the time designated.

I agree to conduct this study in accordance with the declaration of Helsinki and its amendments, the Tri-Council Policy Statement: Ethical Conduct for Research Involving Humans (TCPS-2), International Council for Harmonisation (ICH) Good Clinical Practice Guidelines (GCP) and applicable regulations and laws. I will obtain the approval of a Research Ethics Board for this protocol prior to its implementation.

I have read this protocol and agree that it contains all the necessary details for carrying out this study. I will conduct the study as outlined herein and will complete this study within the time designated.

I will provide copies of the protocol and all pertinent information to all individuals responsible to me who assist in the conduct of this study. I will discuss this material with them to ensure that they are fully informed and trained regarding the study drug(s), the conduct, and the obligations of confidentiality as per the Canadian Privacy Act, The Personal Information Protection and Electronic Documents Act (“PIPEDA”) and the relevant HealthCare Privacy Legislations.

I confirm that I will conduct this clinical trial in compliance with the International Council for Harmonisation Good Clinical Practice Guideline (ICH-GCP E6), the Tri-Council Policy Statement: Ethical Conduct for Research Involving Humans (TCPS-2), applicable Health Canada regulations, the Declaration of Helsinki, the Protocol as approved, and all applicable local and study specific standard operating procedures (SOPs).

| **Site Investigator Name** | **Signature** | **Date** |
| --- | --- | --- |

**Summary**

This is a sub-protocol of the Canadian Adaptive Platform Trial of Treatments for COVID in Community Settings (CanTreatCOVID). For more information on CanTreatCOVID, visit [www.CanTreatCOVID.org](http://www.CanTreatCOVID.org).

Participants meeting the platform, and this sub-protocol inclusion and exclusion criteria will be randomized to receive the following arms:

1. Usual care plus combination antioxidant therapy (comprising of selenium, zinc, lycopene, and vitamin C) QD for 10 days
2. Usual care (i.e., supportive care and symptom relief)

The primary and secondary outcomes are the same as those in the master protocol. CanTreatCOVID uses numerous approaches to recruit, including a multi-faceted public communication strategy and outreach through primary care, out-patient clinics, and emergency departments.

10	ANALYSIS	24[11	REFERENCES	26](#_Toc158985482)

### 1 ABBREVIATIONS

| ACE2 | Angiotensin Converting Enzyme-2 |
| --- | --- |
| AE | Adverse Event |
| CanTreatCOVID | Canadian Adaptive Platform Trial of Treatments for COVID in Community Settings |
| CKD | Chronic Kidney Disease |
| DSMC | Data Safety & Monitoring Committee |
| dNHBE | Differentiated human bronchial epithelial |
| eGFR | Estimated Glomerular Filtration Rate |
| ISSP | Intervention-Specific Sub-Protocol |
| PSSP | Province-Specific Sub-Protocol |
| RAR | Response Adaptive Randomizations |
| SAP | Statistical Analysis Plan |
| SAE | Serious Adverse Event |

### 2 PROTOCOL STRUCURE

The CanTreatCOVID protocol structure, which includes an overarching Master Protocol and intervention-specific sub-protocols (ISSP) is different to that used for conventional trials because this trial is highly adaptive, and the description of these adaptations is better understood and specified using a sub-protocol design. While all adaptations are pre-specified and approved by research ethics prior to implementation, the use of sub-protocols is designed to allow the trial to evolve over time, for example by the introduction or removal of interventions and/or geographical regions.

The protocol structure has multiple elements. In brief, these include a Master Protocol (overview and design features of the study), a Statistical Analysis Plan (SAP; details of the current statistical analysis plan and models), and multiple ISSPs (detailing the individual interventions currently being studied in the trial).

The Master Protocol contains all information that is generic to the trial, irrespective of the provincial location in which the trial is conducted and interventions that are being tested. The Master Protocol may be amended, but it is anticipated that such amendments will be infrequent.

The Master Protocol does not contain information about the intervention(s) included in the trial because one of the trial adaptations is that interventions will change over time. Information about interventions is covered in each ISSP. These ISSPs are anticipated to change over time, with removal, addition, or revision of elements within. Each modification to a ISSP will be subject to an ethics application for approval.

The Master Protocol does not contain detailed information about statistical analyses or simulations, because the analysis model will change overtime in accordance with trial adaptations, however this information is contained in the SAP.

The Master Protocol also does not contain information that is specific to a particular province in which the trial is conducted, as the locations that participate in the trial are also anticipated to change over time. Information that is specific to each province that conducts the trial is contained within a province-specific sub-protocol (PSSP). This includes information related to local management, governance, and ethical and regulatory aspects. It is planned that, within each province, only that province’s PSSP, and any subsequent modifications, will be submitted for ethical review in that province.

### 3 SUB-PROTOCOL SPECIFIC APPENDIX VERSION

The version of the Antioxidant Therapy x 10 days-specific sub-protocol is in this document’s footer and on the cover page.

#### 3.1 Version History

| Version 1.0 | Approved by Unity Health Toronto on: **PENDING** |
| --- | --- |
| Version 2.0 | Approved by Unity Health Toronto on: **PENDING** |
| Version 3.0 | Approved by Unity Health Toronto on: **PENDING** |

### 4 BACKGROUND AND RATIONALE

#### 4.1 Sub-Protocol Definition

This is a sub-protocol within the CanTreatCOVID adaptive platform trial to compare the clinical and cost-effectiveness of a combination antioxidant therapy (comprising of selenium 300ug, zinc 40mg, lycopene 45mg, and vitamin C 1.5g) x 10 days to usual care or other interventions (i.e. Paxlovid™ x 5 days) among non-hospitalized patients with mild to moderate SARS-CoV-2.

#### 4.2 Sub-Protocol-Specific Background

##### 4.2.1 Potential Mechanism of Action

Viral infections induce oxidative stress by producing reactive oxygen species in host cells, which, if not counterbalanced by antioxidant defense mechanisms, can lead to oxidative stress and the emergence of more virulent strains.^3^ The depletion of antiviral defenses and elevated production of inflammatory cytokines have been identified as key factors in SARS-CoV-2 infection. The resulting cytokine storm with high levels of IL-6 and TNF-α produces a critical clinical state, hospitalization, ventilator use and risk of death.

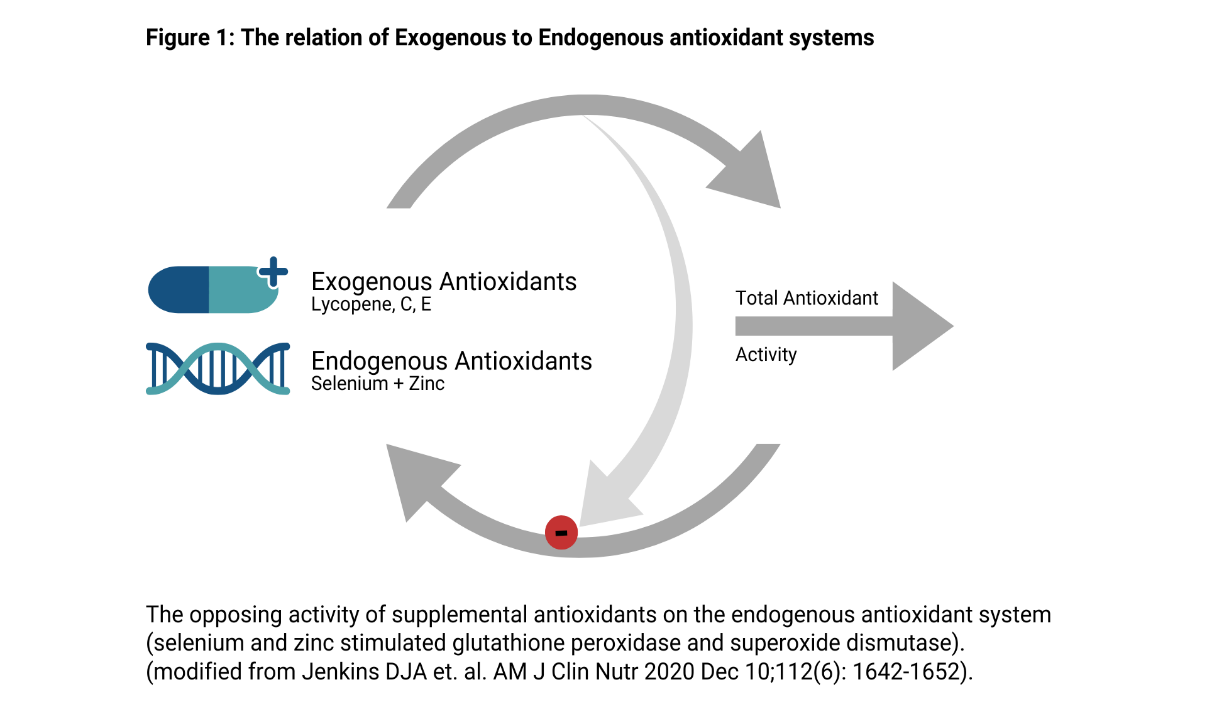
We believe that antioxidants will reduce the prooxidant tissue damage associated with the response to viral infections, providing that both the exogenous and endogenous systems are enhanced. To clarify, our antioxidant defense system includes endogenous (enzymatic and non-enzymatic) antioxidants such as superoxide dismutase, glutathione peroxidase and glutathione, among others and exogenous antioxidants such as vitamin C, vitamin E, and carotenoids, with the diet being the main source.^31^ In previous studies on the effects of antioxidants the focus has been on supplementation with exogenous antioxidants, including vitamin C and E and lycopene, given in combination or singly. Since the total body level of the antioxidant system is maintained in homeostasis, an increase in exogenous antioxidants is generally accompanied by a compensatory reduction of the endogenous system (**Figure 1**)^32^ that minimizes the increase in total antioxidant activity. We therefore propose to provide a powerful exogenous antioxidant in the form of lycopene and vitamin C, that has been shown to reduce the severity of the respiratory viral infections.^1^ Concurrently, selenium and zinc will be provided to strengthen the endogenous system by enhancing the synthesis of glutathione peroxidase and superoxide dismutase, essential elements of the body’s intracellular defense against oxidative stress.

##### 4.2.2 Current Evidence for Potential Benefit(s) of each antioxidant included in the combined antioxidant therapy in SARS-CoV-2 Infection

Selenium

Selenium is an essential structural component of several selenium-dependent enzymes such as glutathione peroxidase, which support the body’s defense mechanism.^2^ Selenium is essential for the proper functioning of both innate and adaptive immunity, including T and B lymphocyte function as well as antibody production, which ultimately impacts susceptibility to infections.^3^ Selenium is also involved in promoting the transcription of anti-inflammatory selenoenzymes, including glutathione peroxidase, as well as reducing the expression of pro-inflammatory inducible enzymes, pro-inflammatory cytokines (such as IL-1, IL-2, IL-6, TNF-α), and chemokines.^4^

In a systematic review of 11 studies, 9 demonstrated negative associations of disease severity and serum selenium levels and 2 showed no effect. Notably, one study demonstrated that renal excretion of selenium was increased with the severity of the disease.^5^ A few randomized controlled trials have also demonstrated the potential benefits of selenium supplementation in improving the immune response against COVID. A randomized, double-blind, placebo-controlled trial of 72 volunteers showed that a combination of active ingredients containing selenium (100 μg/day) increased levels of CD4+ T, CD3+ T, and CD8+ T lymphocytes after the booster dose of the COVID-19 vaccine, with an increase in serum selenium level and no adverse events reported.^6^ Another multicenter, randomized, double-blind, placebo-controlled trial investigated the effects of a nutritional supplement containing selenium (40 μg/day) and other ingredients as an adjunct therapy for COVID-19 hospitalized patients, demonstrating that selenium supplementation reduced the duration of hospital stay and time required to become RT-PCR negative.^7^

Multiple mechanisms involving selenium and selenoproteins at both cellular and viral levels have been implicated in influencing the pathogenicity, severity, and duration of respiratory symptoms, as well as recovery and mortality rates associated with COVID **(Figure 2)**.^8^

**Figure 2:**

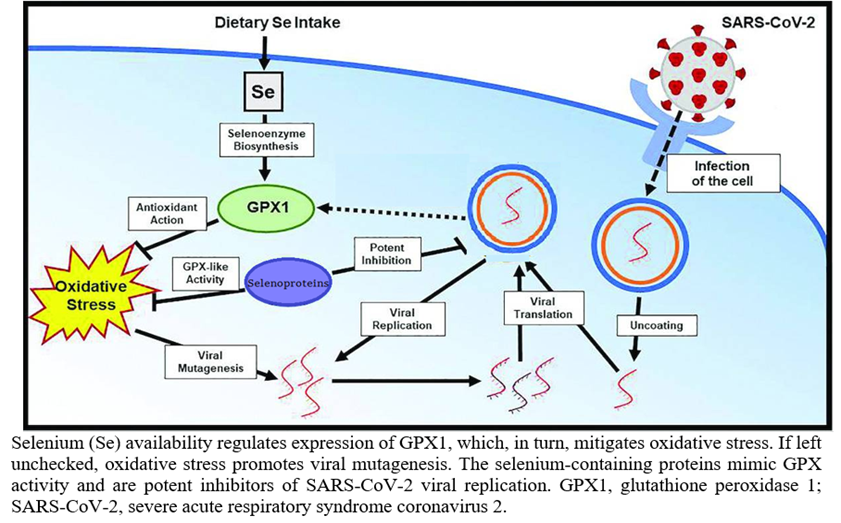

Zinc

Adequate levels of zinc are essential in supporting the endogenous antioxidant system, as it is a key component of superoxide dismutase (SOD) and metallothionein, which are involved in the antioxidant process.^9^ Zinc is also an important component of the Cu, Zn-SOD isoform, responsible for dismutation of superoxide radicals (O^2-^) into hydrogen peroxide (H_2_O_2_). During viral infection, oxidative stress releases zinc from metallothionein to quench free radicals.^10^ Zinc has been shown to inhibit the replication of SARS, polio, and influenza viruses *in vitro*, likely through the formation of zinc ionophores.^11^

In 2015, a Cochrane review of 18 trials on the common cold (16 therapeutic trials, 1387 participants, plus 2 prevention trials, 394 participants) demonstrated that zinc supplementation resulted in a reduction of duration of symptoms by 1.03 days (95% CI 1.72 to 0.34) (p=0.003) (I2=89%), with a markedly reduced school absence (p=0.00003) and reduced antibiotic use (p<0.000001).^12^ A meta-analysis of 3 trials and 2 cohort studies reported that zinc supplementation was associated with a significantly lower risk of mortality compared to the control group, with a pooled odds ratio (OR) of 0.57 (95% confidence interval [CI]: 0.43 to 0.77; p < 0.001).^13^ A recent multicenter RCT on 190 outpatients and 280 hospitalized patients provided further evidence for the beneficial effects of zinc supplementation against COVID morbidity and mortality.^14^ The trial showed that zinc supplementation (50 mg/day for 15 days) reduced mortality (OR: 0.68; 95% CI 0.34 to 1.35) and ICU admission rates (OR: 0.43; 95% CI 0.21 to 0.87). Additionally, the duration of COVID-19 symptoms decreased following zinc treatment compared to placebo in outpatients, with a difference of 1.9 days (95% CI 0.62 to 2.6). Consistent results were observed in prespecified subgroups of patients aged <65 years, those with comorbidity, and those who needed oxygen therapy at baseline.

Lycopene

Lycopene can scavenge oxygen radicals, reduce oxidative stress, and prevent ROS generation, and has been regarded as the most effective ^1^O_2_ scavenger *in vitro* of all the carotenoids.^15^ The major complication of SARS-CoV-2 infection, the acute respiratory distress syndrome (ARDS), is due to a variety of mechanisms such as cytokine storm and neutrophil activation.^16,17^

Lycopene has been shown to provide protection against airway inflammation induced by rhinovirus, the most common cause of the common cold in adults and children, by reducing expression of pro-inflammatory markers in airway epithelial cells following rhinovirus-1B infection (24% and 31% reduction in IL-6, IP-10; p<0.05). In addition, lycopene reduced Rhinovirus-1B replication by 85% in airway epithelial cells (p=0.025).^18^

Studies have demonstrated that SARS-CoV-2 infection in airway epithelial cells results in elevated levels of IL-6 expression at 24 hours post-infection, which can be inhibited with remdesivir treatment.^19^ Interestingly, lycopene's ability to reduce IL-6 in response to rhinovirus, a virus that shows similar effects on inflammatory cytokines in epithelial cells as SARS-CoV-2 infection, appears to be comparable to that of remdesivir. Furthermore, in a previous randomized clinical trial, the administration of tomato juice (45 mg Lycopene/day) and extract (45 mg Lycopene/day) for 7 days resulted in a reduction of neutrophil elastase activity in the airways.^20^ This finding is particularly relevant considering the growing evidence suggesting the role of neutrophil elastase in the pathogenesis of respiratory infectious diseases, including SARS-CoV-2.^21^ Indeed, studies have shown that levels of both neutrophil elastase and IL-6 were significantly higher in deceased COVID-19 patients compared to recovered patients.^21^

Lycopene supplementation also tended to be positively associated with pulmonary function. Short-term (1 week) supplementation with tomato extract (containing 30 mg lycopene/ day) led to a reduction in exercise-induced bronchoconstriction (post-exercise reduction in FEV1: Lycopene intervention: 14% vs. Placebo: 26.5%; p<0.05).^22^ Recent studies demonstrated that long-term pulmonary impairment affects approximately 3%–5% of individuals following COVID-19, particularly those over the age of 50.^23^ After 100 days of COVID-19 diagnosis, nearly one-third of patients reported experiencing dyspnea and impaired pulmonary function, while two-thirds showed abnormal pulmonary imaging findings.^24^ Given the potential for long-term pulmonary impairment following COVID-19, lycopene supplementation may be beneficial in mitigating these effects, particularly in those over the age of 50 who are at a higher risk.

Vitamin C

Several RCTs have demonstrated that regular administration of vitamin C at a dosage of greater than 1 g/day can reduce the duration of respiratory viral infections by 8% in adults and 18% in children.^25^ Similarly, a more recent meta-analysis on severity and duration of symptoms indicated that therapeutic levels of supplementation (≥0.2 g/day) may have benefits for symptom duration.^1^ As vitamin C has been shown to have non-specific effects on a diverse group of respiratory viruses, it seems plausible that it could also have an impact on SARS-CoV-2.

In a recent survey of 18 COVID-19 patients, 17 had undetectable vitamin C levels, while the remaining patient had a very low level.^26^ Similarly, another study found that COVID-19 patients had low vitamin C plasma levels, with non-survivors having only half the plasma level of survivors.^27^ A meta-analysis of 12 trials involving 1766 patients revealed that vitamin C administration reduced the length of ICU stay by an average of 7.8% (95% CI: 4.2% to 11.2%; p = 0.00003).^28^ Additionally, orally administered vitamin C at a dosage of 1-3 g/day (weighted mean 2.0 g/day) resulted in an 8.6% reduction in ICU stay length (p = 0.003).28 Moreover, in three trials where mechanical ventilation was required for more than 24 hours, vitamin C administration shortened the duration of mechanical ventilation by 18.2% (95% CI 7.7% to 27%; p = 0.001).28 Furthermore, two studies have reported significant reductions in mortality rates with the use of vitamin C in critically ill patients with sepsis and acute respiratory distress syndrome (ARDS). Zabet et al. found that vitamin C reduced mortality by 78% (P = 0.01; based on 2/14 vs. 9/14) in 28 sepsis patients,^29^ while Fowler et al. reported a 35% reduction in mortality (P = 0.01; based on 25/84 vs. 38/83) in 167 patients with sepsis and ARDS.^30^

### 5 SUB-PROTOCOL OBJECTIVES

The objective of this sub-protocol is to determine the clinical- and cost-effectiveness of antioxidant therapy x 10 days among non-hospitalized patients with mild to moderate SARS-CoV-2 infection.

We hypothesize that the treatment effect of the combination antioxidant therapy x 10 days is different depending on age and vaccination strata status.

This is one of the intervention arms investigated in CanTreatCOVID, and participants are randomized to the arms for which they are eligible.

### 6 TRIAL DESIGN

The sub-protocol will be conducted as part of the CanTreatCOVID trial. Treatment arm allocation will occur on the day of enrollment using an interactive web-based system, as described in the Master Protocol. Briefly, participants will be allocated to a trial arm using fixed, equal randomization ratios corresponding to the number of eligible arms in the trial. Participants will be stratified based on age and random sized permuted blocks will be used.

#### 6.1 Population

CanTreatCOVID enrolls non-hospitalized patients with mild to moderate SARS-CoV-2 infection, regardless of their COVID vaccination status.

#### 6.2 Eligibility Criteria

Participants are eligible for this sub-protocol if they meet all of the platform-level inclusion and none of the platform-level exclusion criteria as specified in the CanTreatCOVID Master Protocol.

Participants otherwise eligible for CanTreatCOVID may have conditions that exclude them from the current sub-protocol. These are described below.

##### 6.2.1 Sub-Protocol Inclusion Criteria

As per the Master Protocol, except for age limit as participants would have to be 19+ years old.

##### 6.2.2 Sub-Protocol Exclusion Criteria

Participants will be excluded from this sub-protocol if the participant:

- Has a known or suspected pregnancy.
- Is breastfeeding.
- Is of childbearing potential and is not willing to use a highly effective contraceptive.
- Has allergy or intolerance to selenium, zinc, lycopene, vitamin C, ascorbyl palmitate, hypromellose, microcrystalline cellulose, or sodium stearyl fumarate
- Is taking warfarin as a preventive measure.
- Has advanced chronic kidney disease (CKD stage 3: eGFR ≥30 to <60 mL/min, and severe renal impairment (eGFR <30 ml/min, CKD stage 4-5)
- Has liver disease awaiting transplantation.
- Has a history of calcium oxalate kidney stones.
- Has a diagnosis of head and neck cancer within the past 5 years
- Has medical history of non-melanoma skin cancer
- Is taking selenium (≥300ug/day), zinc (≥40mg/day), lycopene (≥45mg/day), and vitamin C (≥1500mg/day) supplements at baseline
- Is consuming omega-3 fatty acids at baseline and is unwilling to stop during the intervention period.

To confirm that the participant meets the criteria defined above, information will be elicited through a direct discussion between the participant and a medically qualified prescribing pharmacist. Those assessing eligibility will take a relevant drug history and obtain a complete medication list from the participant’s usual pharmacy. If after reviewing this list and discussion with the patient, the recruiting health care professional considers the potential participant eligible, the participant may then be randomised to the Antioxidant Therapy arm.

#### 6.3 Interventions

Participants will be randomly allocated to one of the following trial arms:

1. Combination antioxidant therapy x 10 days
2. Usual Care (i.e., supportive care and symptom relief) (details described in the master protocol)
3. Additional treatments may be added to the trial in the future, expanding the intervention options.

CanTreatCOVID is an open-label trial so participants will be aware of which trial arm they have been allocated to. If a participant is allocated to any of the intervention arms, the respective medication will be shipped to their residence immediately after their eligibility is confirmed.

##### 6.3.1 Combined antioxidant therapy x 10 days

*Dosing and Duration of Treatment*

The daily dosage level will include 300 ug of selenium, 40 mg of zinc, 45 mg of lycopene, and 1.5 g of vitamin C, which will be given as three capsules taken once per day for 10 days. Participants will be advised to complete the full 10-day treatment course.

The 10-day intervention course should be initiated as soon as possible after a diagnosis of SARS-CoV-2 has been made, and within 5 days of symptom onset.

Adherence to trial medication will be assessed by self-report. Participants will be asked not to take any natural health products, including other vitamin/mineral supplements, or any supplement providing any additional dose of selenium, vitamin C, lycopene, or zinc, for the duration of the intervention.

*Dosing Considerations*

The participant can take the three capsules composed of the antioxidants only once at any time of the day. Antioxidant supplementation should be taken with food, a few hours before or after taking other medications or natural health products. The capsules should be swallowed whole and not chewed, broken, or crushed.

However, if the participants missed to take a dose for an entire day, then the participant must skip that dose and continue with their scheduled doses on their respective dates. The participant should not double the dose to make up for a missed dose.

If a participant overdoses with the antioxidant therapy, they will be advised to get in touch with their primary care provider or local emergency department, if required.

If a patient requires hospitalization due to severe or critical COVID-19 after starting treatment with the antioxidant therapy, the patient should complete the full 10-day treatment course at the discretion of their healthcare provider.

*Renal Failure*

No dose adjustment is necessary for patients with mild renal impairment. Patients with moderate renal impairment (eGFR ≥30 to <60 mL/min, CKD stage 3) and severe renal impairment (eGFR <30 ml/min, CKD stage 4-5) are not recommended for the antioxidant therapy arm and will not be eligible for randomization to the antioxidant therapy arm.

*Hepatic impairment*

No dose adjustment is required for patients with mild to severe hepatic impairment. Only patients with hepatic impairment that are awaiting liver transplantation are not recommended for the antioxidant therapy arm and are not eligible for randomization to the antioxidant therapy arm.

*Excipients*

There are no excipients in the antioxidant formulation; however, the capsules themselves are gelatin-based.

*Concomitant medications*

The antioxidant formulation is a custom-made formulation prepared by Advanced Orthomolecular Research Inc. (AOR) based in Alberta. All four antioxidants are combined in an oral dosage form of three capsules per day, without any added excipients. Drug-drug interactions should be managed as per the published drug-drug interactions of each individual antioxidant (see Section 8.1.2 Risk of drug interactions)

During the 10 days of treatment, participants who are taking the investigational product as part of the trial will be advised to inform their health care providers (i.e. participant information sheet – Appendix 3, wallet card – Appendix 4) about their participation.

#### 6.4 Concomitant Care

All treatment that is not specified by assignment within the current trial will be determined by the participant’s usual health care provider.

#### 6.5 Endpoints and Outcomes

##### 6.5.1 Primary Outcomes

As per the Master Protocol.

##### 6.5.2 Secondary Outcomes

As per the Master Protocol.

### 7 TRIAL CONDUCT

#### 7.1 Sub-Protocol-Specific Data Collection

##### 7.1.1 Clinical Data Collection

An additional safety call will be made on Day 6 (Appendix 6) for participants randomized to the Antioxidant Therapy arm. The purpose of the day 6 safety call is to detect any side-effects of the antioxidant therapy and to enable investigators to suggest changes to participants’ medication including stopping where required. We will obtain two 24-hour dietary recalls (24-HDR) from participants randomized to antioxidant therapy and also usual care: one at the study's baseline and another at the end of intervention (10-day). 24-hours recall will be conducted by telephone by trained interviewers. During the 24-HDR, each subject will be asked to recall and describe in detail, all types and amounts of foods and beverages consumed in the previous 24 hours on two separate occasions, a weekday and a weekend day. Weekend days included Saturday and Sunday to capture food and alcohol consumption patterns which may be different from those on weekdays (Monday to Friday).^41,42,43^ The 24-hour period specified for the dietary recall is defined as the 24 consecutive hours between midnight on day one and midnight on the following day. To assist in estimating portion sizes of consumed foods, participants will be encouraged to view a measuring cup and measuring spoons as they completed their 24-HDR by telephone. At the end of this study, there will be a total of four completed 24-HDRs for each participant. At baseline, participants will also be asked to complete a 169-item food frequency questionnaire (FFQ) validated for the Canadian population.^44^ The FFQ requires participants to recall the frequency of consumption for each food item (per day, per week, per month, or rarely/never) over the past 12 months. Additionally, participants must indicate the number of months during the year each food was consumed to account for seasonal variations in intake. Portion size options will be provided using standard measuring units (e.g., cups, tablespoons, slices) or by referring to photographs representing small, medium, and large portion sizes for some food items.

Results from 24-hour dietary recalls (24HDR) and food frequency questionnaires (FFQs) will be used to collect data on the quantity of consumption of known antioxidant-rich foods and supplements. We will then calculate the Dietary Antioxidant Score (DAS) for each participant both before and after the intervention. DAS is an indirect measure of participants’ antioxidant intake.^38, 39^

#### 7.2 Criteria for Discontinuation

Refer to the CanTreatCOVID Master Protocol for criteria for discontinuation of participation in the trial.

The study doctor may stop the participant’s participation in the study early, and without their consent, for a variety of reasons:

- The participant is no longer considered eligible for the study.
- New information shows that the study group activity (standard care, or a specific medication) is no longer in the participant’s best interest.
- The study doctor no longer feels this is the best option for the participant.
- The participant is unable to tolerate the study medication.

#### 7.3 Blinding

##### 7.3.1 Blinding

Combined Antioxidant Therapy x 10 days will be administered on an open-label basis.

##### 7.3.2 Unblinding

Not relevant.

#### 7.4 Statistical Analysis

Primary analysis will be conducted using the intention-to-treat principle whereby patients with protocol violations will be analyzed in the arm to which they were allocated. To determine the primary outcome, patient-reported data will be relied upon. Several attempts to follow up with patients and provide reminders to take the study supplements will be made during the first contact to minimize risk of bias from non-adherence.

Tabulations will be produced for appropriate demographic, baseline, efficacy, and safety parameters. For categorical variables, summary tabulations of the number and percentage within each category (with a category for missing data) of the parameter will be presented. For continuous variables, the mean, median, standard deviation, minimum and maximum values will be presented. For the primary endpoint, Bayesian analysis with covariates adjustment will be performed. Randomized group and age will be included as covariates. Summary statistics of log odds ratio for each pairwise treatment arm comparison with the usual care group will be presented, along with their corresponding 95% Bayesian credible intervals. For the secondary endpoints that are ordinal or binary, logistic regression, ordinal logistic regression, and linear regression will be used. Randomized group and age will be included as covariates. Where logistic regression and ordinal logistic regression are used, the odds ratios will be reported for each pairwise treatment arm comparison with the usual care group, along with their respective 95% confidence interval. Where linear regression is used, the mean and standard deviation will be reported in each treatment arm. The adjusted difference in means, along with the 95% confidence interval for each pairwise treatment arm comparison with the usual care group, will be reported. For the secondary time-to-event type endpoints, Cox’s proportional hazards model will be applied. Randomized group and age will be included as covariates. Summary statistics of log hazards ratio for each pairwise treatment arm comparison with the usual care group will be presented, along with their corresponding 95% confidence intervals. In addition, time to recovery data will be summarized using Kaplan-Meier estimates.

***Sex and gender analysis***: Moreover, there will be an investigation of the intersectionality of sex and gender with other social determinants of health, such as income, race, ethnicity, food security and rurality.

***Planned sub-group analysis:*** Subgroup analyses will be performed on several populations, including age, sex, gender, body mass index (BMI), income, race and ethnicity, rurality, medical conditions (multimorbidity and immunocompromised), number of vaccine doses received (< 2 vs ≥2), and place of recruitment (emergency departments vs other settings). There will also be an examination of the moderating effects of concomitant medications, such as Paxlovid and corticosteroids. For each subgroup, calculations of the model-based estimates of the difference in rates of emergency visits, death, and hospitalization between the treatment arms and the usual care, accompanied by 95% Bayesian credible intervals computed using the primary analysis model, will take place.

### 8 ETHICAL CONSIDERATIONS

#### 8.1 Risks

##### 8.1.1  Adverse events

The doses selected for the combined antioxidant therapy are below the Tolerable Upper Intake Level (UL) to ensure long-term safety and to provide maximum potential benefits. The UL doses for adults are 1.8g for vitamin C, 400ug for selenium, and 40mg for zinc, while lycopene has no UL.^52^ Adverse events and serious adverse events may be collected through the daily diary, communicated on study surveys or to research staff directly, or communicated through the toll-free study hotline maintained by study staff between 8 AM to 6 PM ET. Analyses of treatment-emergent adverse events will be performed, and the results will be summarized by subject incidence rates. Each subject will only be counted once in any tabulation for a given adverse event. No formal hypothesis-testing analysis of adverse events incidence rates will be performed, and all adverse events will be listed in subject data listings. By-subject listings will be provided for subject deaths, serious adverse events, and adverse events leading to withdrawal.

In the following section, we will describe possible adverse events in patients following a 10-day antioxidant supplementation regimen.

- **Selenium:** Excessive selenium intake can lead to selenosis, characterized by gastrointestinal discomfort, hair loss, fatigue, irritability, and mild nerve damage. The narrow therapeutic-to-toxic ratio of selenium necessitates careful monitoring for signs of overdose.^33^
- **Zinc:** High zinc doses can impede copper absorption, leading to potential copper deficiency. Common side effects include nausea, vomiting, and stomach pain.^34^
- **Lycopene:** Lycopene is usually well-tolerated, but excessive intake might cause gastrointestinal issues or skin discoloration. Allergic reactions, though rare, should be considered in patients with unexplained symptoms.^35^
- **Vitamin C:** High vitamin C doses can lead to gastrointestinal disturbances and may contribute to kidney stone formation in susceptible individuals.^36^

##### 8.1.2 Risk of drug interactions

Interactions with Vitamin C: ^37^

| Concomitant Medication | Management (Rationale) |
| --- | --- |
| Aluminum Hydroxide | Avoid coadministration (Lexicomp Risk Rating D: increased systemic absorption of Aluminum) |
| Deferoxamine | Avoid coadministration (Lexicomp Risk Rating D: Vitamin C doses greater than 200mg/day may enhance toxicity of Deferoxamine) |
| Amphetamines (amphetamine, dextroamphetamine, lisdexamfetamine, methadmphetamine) | Monitor therapy with concomitant medication (Lexicomp Risk Rating C: gastrointestinal acidifying agents may decrease systemic absorption of Amphetamines) |
| Cyclosporine | Monitor therapy with concomitant medication (Lexicomp Risk Rating C: decreased systemic absorption of Cyclosporine) |
| Warfarin | Monitor therapy with concomitant medication (Lexicomp Risk Rating C: decreased anticoagulant effects and will require additional anticoagulation monitoring ) |

Interactions with Selenium: ^37^

| Concomitant Medication | Management (Rationale) |
| --- | --- |
| Baloxavir Marboxil | Avoid coadministration (Lexicomp Risk Rating X: polyvalent cation containing products may decrease the serum concentration of Baloxavir Marboxil) |
| Unithiol | Avoid coadministration (Lexicomp Risk Rating X: Unithiol may diminish the therapeutic effects of polyvalent cation containing products) |
| Bictegravir | Separate administration by taking Bictegravir 2 hours before or 6 hours after Selenium (Lexicomp Risk Rating D: polyvalent cation containing products may decrease the serum concentration of Bictegravir) |
| Bisphosphonate derivatives | Separate administration as follows:  Etidronate: 2 hours (before or after Selenium)  Ibandronate: 1 hour (before or after Selenium)  Alendronate & risedronate: 30minutes (before or after Selenium) (Lexicomp Risk Rating D: polyvalent cation containing products may decrease the serum concentration of Bisphosphonate derivatives) |
| Cabotegravir | Separate administration by taking Selenium either 2 hours before or 4 hours after Cabotegravir (Lexicomp Risk Rating D: polyvalent cation containing products may decrease the serum concentration of oral Cabotegravir) |
| Deferiprone | Separate administration by taking Selenium at least 4 hours before or after Deferiprone (Lexicomp Risk Rating D: polyvalent cation containing products may decrease the serum concentration of oral Deferiprone) |
| Dolutegravir | Separate administration by taking Dolutegravir either 2 hours before or 6 hours after Selenium (Lexicomp Risk Rating D: polyvalent cation containing products may decrease the serum concentration of oral Dolutegravir) |
| Eltrombopag | Separate administration by taking Eltrombopag either 2 hours before or 4 hours after Selenium (Lexicomp Risk Rating D: polyvalent cation containing products may decrease the serum concentration of oral Eltrombopag) |
| Elvitegravir | Separate administration by taking Elvitegravir either 2 hours before or 6 hours after Selenium (Lexicomp Risk Rating D: polyvalent cation containing products may decrease the serum concentration of oral Elvitegravir) |
| Penicillamine | Separate administration by taking Selenium at least 1 hour before or after Penicillamine (Lexicomp Risk Rating D: polyvalent cation containing products may decrease the serum concentration of oral Penicillamine) |
| Raltegravir | Separate administration by taking Raltegravir either 2 hours before or 6 hours after Selenium (Lexicomp Risk Rating D: polyvalent cation containing products may decrease the serum concentration of oral Raltegravir) |
| Roxadustat | Administer Roxadustat at least 1 hour after taking Selenium (Lexicomp Risk Rating D: polyvalent cation containing products may decrease the serum concentration of oral Roxadustat) |
| Trientine | Separate administration by taking Trientine at least 1 hour before or 1 to 2 hours after Selenium (Lexicomp Risk Rating D: polyvalent cation containing products may decrease the serum concentration of oral Trientine) |

Interactions with Zinc: ^37^

| Concomitant Medication | Management (Rationale) |
| --- | --- |
| Baloxavir Marboxil | Avoid coadministration (Lexicomp Risk Rating X: polyvalent cation containing products may decrease the serum concentration of Baloxavir Marboxil) |
| Levonadifloxacin | Avoid coadministration (Lexicomp Risk Rating X: polyvalent cation containing products may decrease the serum concentration of Levonadifloxacin) |
| Unithiol | Avoid coadministration (Lexicomp Risk Rating X: Unithiol may diminish the therapeutic effects of polyvalent cation containing products) |
| Bictegravir | Separate administration by taking Bictegravir 2 hours before or 6 hours after Zinc (Lexicomp Risk Rating D: polyvalent cation containing products may decrease the serum concentration of Bictegravir) |
| Bisphosphonate derivatives | Separate administration as follows:  Etidronate: 2 hours (before or after Zinc)  Ibandronate: 1 hour (before or after Zinc)  Alendronate & risedronate: 30minutes (before or after Zinc) (Lexicomp Risk Rating D: polyvalent cation containing products may decrease the serum concentration of Bisphosphonate derivatives) |
| Cabotegravir | Separate administration by taking Zinc either 2 hours before or 4 hours after Cabotegravir (Lexicomp Risk Rating D: polyvalent cation containing products may decrease the serum concentration of oral Cabotegravir) |
| Deferiprone | Separate administration by taking Zinc at least 4 hours before or after Deferiprone (Lexicomp Risk Rating D: polyvalent cation containing products may decrease the serum concentration of oral Deferiprone) |
| Dolutegravir | Separate administration by taking Dolutegravir either 2 hours before or 6 hours after Zinc (Lexicomp Risk Rating D: polyvalent cation containing products may decrease the serum concentration of oral Dolutegravir) |
| Eltrombopag | Separate administration by taking Eltrombopag either 2 hours before or 4 hours after Zinc (Lexicomp Risk Rating D: polyvalent cation containing products may decrease the serum concentration of oral Eltrombopag) |
| Elvitegravir | Separate administration by taking Elvitegravir either 2 hours before or 6 hours after Zinc (Lexicomp Risk Rating D: polyvalent cation containing products may decrease the serum concentration of oral Elvitegravir) |
| Penicillamine | Separate administration by taking Zinc at least 1 hour before or after Penicillamine (Lexicomp Risk Rating D: polyvalent cation containing products may decrease the serum concentration of oral Penicillamine) |
| Quinolones | Separate administration as follows:  Moxifloxacin: 4 hours before or 8 hours after Zinc  Ciprofloxacin: 2 hours before or 6 hours after Zinc  Lomefloxacin: 2 hours before or 4 hours after Zinc  Gemifloxacin: 2 hours before or 3 hours after Zinc  Levofloxacin, Norfloxacin, Ofloxacin, Nalidixic Acid: 2 hours before or after Zinc |
| Raltegravir | Separate administration by taking Raltegravir either 2 hours before or 6 hours after Zinc (Lexicomp Risk Rating D: polyvalent cation containing products may decrease the serum concentration of oral Raltegravir) |
| Roxadustat | Administer Roxadustat at least 1 hour after taking Zinc (Lexicomp Risk Rating D: polyvalent cation containing products may decrease the serum concentration of oral Roxadustat) |
| Specific Tetracyclines (only applies to Demeclocycline, Minocycline, Tetracycline) | Separate administration from Zinc by at least 2 hours (Lexicomp Risk Rating D: zinc salts may decrease the absorption of the specific Tetracyclines) |
| Trientine | Separate administration by taking Trientine at least 1 hour before or 1 to 2 hours after Zinc (Lexicomp Risk Rating D: polyvalent cation containing products may decrease the serum concentration of oral Trientine) |

Interactions with Lycopene: ^37^

No relevant drug-drug interactions that require active management.

##### 8.1.3 Risks in pregnancy and during breastfeeding

There is no human data on the effect of the combined antioxidant therapy on pregnancy or in breastfeeding. The summary of product characteristics states that breastfeeding should be discontinued during treatment with the antioxidant therapy and for 14 days after the last dose of the antioxidant therapy. Therefore, to be eligible for randomization to the antioxidant therapy arm, participants are required to use a highly effective method of contraception for the duration of the treatment and 28 days of follow-up. Pregnant and breastfeeding participants will not be eligible.

If a participant decide to participate in the study and if there is any chance they could conceive a child, they will need to use a ‘highly effective’ method of contraception and/or abstinence for 38 days (this number is the total of 10 days of the antioxidant therapy plus 28 days after).

- Please note, no one method of contraception is 100% effective.
- Highly effective methods include:
  - Hormonal contraceptives like combined oral contraceptives, patch, vaginal ring, injectables, and implants.
  - Long-acting reversible contraceptives like an intrauterine device (IUD) or intrauterine system (IUS).

#### 8.2 Benefits

Current antiviral therapies for SARS-CoV-2 infection have limitations, such as potential adverse effects ranging from mild symptoms to severe reactions, and eligibility restrictions for certain patient populations, that limit their effectiveness and accessibility. These limitations underscore the need for alternative and safe therapeutic approaches for COVID. Antioxidant therapy offers a promising solution, with the potential to provide similar results to current outpatient therapeutics, without the risk of side effects, and at a lower cost. Several systematic reviews have suggested that antioxidants may have potential benefits for COVID-related outcomes, including preventing disease progression in COVID-19 patients with higher levels of antioxidants.^38,39^ While it is not reasonable to claim that antioxidants are a solution to the COVID pandemic, emerging evidence suggests that they may have a role in preventing disease progression or supporting therapy in intensive care settings. The physiology, pharmacology, and basic science behind our antioxidants of interest point to potential benefits that warrant further investigation, even if this means focusing on diligent deficiency correction rather than mass supplementation. Moreover, supplementation with these antioxidants, such as vitamin C has been shown to reduce the duration of respiratory infections. Therefore, it is plausible that antioxidant therapy could decrease symptom duration and time to recovery in mild to moderate cases of SARS-CoV-2, similar to the effects of Molnupiravir demonstrated in the PANORAMIC trial^40^, but without requiring the close monitoring necessary for medications such as Molnupiravir.

#### 8.3 Data Safety and Monitoring Committee

The DSMC will receive results reported as Group A (arm 1), Group B (arm 2), etc. Should there be a significant difference in outcomes or adverse events, data will be unblinded to the DSMC chair and then shared with the Steering Committee to reach a unanimous decision with respect to next steps (i.e., continue or stop the trial). Stopping rules are pre-determined and included in the SAP related to efficacy, futility, and safety, and sub-protocol stopping rules.

The DSMC should be aware that the superiority, efficacy, inferiority, futility, or equivalence of different interventions with respect to the co-primary outcomes are possible, and if equivalence is demonstrated, determination of the optimal intervention may be based on secondary outcomes. The DSMC should take into account the public health, as well as clinical significance, of the analyses of this sub-protocol and are empowered to discuss results with relevant international and national public health authorities, with rapid dissemination of results to the larger community being the goal.

#### 8.4 Sub-Protocol Specific Consent Concerns

As endorsed by the World Health Organization, in the absence of evidence of effectiveness of specific treatments for SARS-CoV-2, the use of a usual care control is both appropriate and ethical.

Health care providers will be directed not to refer an individual patient for enrollment if they believe that participation in this sub-protocol is not in the best interests of the patient.

### 9 GOVERNANCE CONCERNS

#### 9.1 Funding of Sub-Protocol

Funding sources for CanTreatCOVID are specified in the Master Protocol and sub-protocol cover page.

#### 9.2 Funding of Sub-Protocol Interventions

The combined antioxidant therapy will be provided by the Advanced Orthomolecular Research (AOR), Calgary, Canada.

#### 9.3 Sub-Protocol-Specific Declarations of Interest

All members of the CanTreatCOVID Canadian COVID-19 Out-Patient Therapeutics Committee, which makes decisions regarding the treatments to be evaluated in this trial are collected and updated on an annual basis.

￼

### 11 APPENDICES

Appendix 1 - Information sheet and consent

Appendix 2 - Initial Contact Script and Screening Form

Appendix 3 - Participant Information Sheet

Appendix 3.1- Standard of Care Information Sheet

Appendix 4 - Wallet Contact Card

Appendix 5 - Product Monogram

Appendix 5.1 - Drugs that Require Adjustment when Co-Administered

Appendix 6 - Follow-up at Day 6

Appendix 7 – FFQ Questionnaire
